# MRI Characterization of Structural Brain Abnormalities in NGLY1 Deficiency

**DOI:** 10.64898/2026.03.30.26348623

**Authors:** Emily L Dennis, Lei Zhu, William F Mueller, Jennifer W Cook, Gracie Morrison, Matt Wilsey, Ryan Dant, Selina Dwight, Kevin J Lee

**Author notes:** Please direct correspondence to: Kevin Lee, PhD. Chief Scientific Officer, Grace Science Foundation.

## Abstract

**Key Points:** *Question:* Is NGLY1 Deficiency associated with consistent alterations in brain structure and do these alterations relate to clinical phenotypes?

*Findings:* In this case series of 11 patients with NGLY1 Deficiency, MRI brain scans revealed significant subcortical volume reductions across patient age groups, particularly in the thalamus, caudate, and putamen. Younger patients (<3 years old) showed extensive cortical disruptions suggestive of altered gyrification, while older patients (>3 years old) demonstrated region-specific cortical thickness changes. These alterations were associated with several clinical variables, including the presence of seizures, dysphagia, gait disturbance, EEG abnormalities, sleep disturbance, and hearing abnormalities.

*Meaning:* Widespread alterations in cortical and subcortical morphology as assessed by MRI highlight the significant impact NGLY1 Deficiency has on brain development. Associations between morphology and clinical variables suggest that these neuroimaging metrics may serve as useful biomarkers of disease progression and in assessing treatment efficacy.

**Importance:** NGLY1 (N-Glycanase 1) Deficiency is an ultra-rare autosomal recessive disorder affecting ∼165 patients worldwide, characterized by developmental delay, hyperkinetic movement disorders, and shortened life expectancy. Despite its severe neurological manifestations, comprehensive neuroimaging characterization has been limited to case reports and small descriptive studies.

**Objective:** To investigate alterations in brain morphology in patients with NGLY1 Deficiency and determine whether these metrics associate with clinical phenotypes.

**Design, Setting, and Participants:** This case series analyzed real-world MRI scans performed on 11 patients with NGLY1 Deficiency between 1999-2023 at sites across the globe. Ages ranged from 2 to 19 years at scan time (5 female, 6 male).

**Exposure:** Molecular diagnosis of NGLY1 Deficiency.

**Main Outcomes and Measures:** Cortical and subcortical morphology, including subcortical volume, and cortical thickness, surface area, volume, and curvature, were measured with 3-dimensional T1-weighted magnetic resonance imaging (MRI) scans. Z-scores were calculated using normative models from CentileBrain for patients >3 years old or custom models for patients <3 years old. Clinical phenotypes were matched to Human Phenotype Ontology codes.

**Results:** 16 scans from 11 patients met quality criteria for analysis. Both age groups (under and over 3 years old) showed significantly reduced subcortical volumes, particularly in bilateral thalamus and putamen. Younger patients demonstrated widespread reductions in cortical surface area, volume, and curvature, indicating altered gyrification patterns. Older patients showed thinner dorsal and thicker ventral cortical regions with limited surface area reductions. Thalamic volume reduction in older patients correlated with gait disturbance, dysphagia, and EEG abnormalities, with additional cortical associations with sleep and hearing abnormalities. Seizure presence in younger patients correlated with altered cortical thickness, surface area, and curvature patterns.

**Conclusions and Relevance:** NGLY1 Deficiency is associated with pervasive alterations in brain development affecting both subcortical and cortical morphology. Age-dependent patterns of cortical alterations indicate disrupted neurodevelopmental trajectories that may reflect impaired neuronal migration and/or altered synaptic pruning. Correlations with clinical variables suggest that these measures may serve as useful biomarkers for tracking disease progression and/or treatment efficacy. These findings provide a comprehensive neuroimaging characterization of NGLY1 Deficiency and establish a foundation for understanding brain structure-function relationships in this ultra-rare disorder.

## Introduction

NGLY1 (*N*-Glycanase 1) Deficiency is an ultra-rare, debilitating autosomal recessive disorder with ∼165 patients identified worldwide to date,^1,2^ although one study reports the true incidence may be closer to 12 births/year in the United States.^3^ It is caused by loss of function mutations in the NGLY1 gene that impair endoplasmic reticulum-associated degradation (ERAD) and the clearance of abnormal glycoproteins within the cytosol.^4^ Patient phenotypes are heterogeneous, varying in prevalence and severity. The core phenotypes can be characterized by severe global developmental delay and/or intellectual disability, hyperkinetic movement disorder, transient elevation of transaminases, (hypo)alacrima, and progressive, diffuse, length-dependent sensorimotor polyneuropathy. Other, more diverse, phenotypes have been described including epilepsy, scoliosis, dysphagia, EEG abnormalities, sleep disturbances, and microcephaly.^4^ A recent natural history study by our group followed 29 patients over a few years and characterized clinical features and disease course, noting motor decline over time.^5^

Neuroimaging findings in NGLY1 Deficiency have been limited to descriptive reports and a single small-scale systematic study. Lam and colleagues collected brain magnetic resonance imaging in 11 patients ranging in age from 3-21 years old, describing delayed myelination in three of four youngest individuals (under 6 years old), cerebral atrophy in six patients (ranging from slight to moderate), and slight cerebellar atrophy in four patients, with atrophy correlating strongly with functional impairment as measured by the Nijmegen Pediatric CDG Severity scale and the Vineland Adaptive Behavior Scale.^6^ Additional case reports have described non-specific findings including delayed myelination, atrophy of the corpus callosum, anterior commissure and/or cerebellum, prominent cisterna magna, and enlarged ventricles.^7–14^ However, comprehensive analyses of brain morphometry have not been reported. Understanding the specific neuroanatomical profile of NGLY1 Deficiency may provide crucial insights into the pathophysiological mechanisms underlying the cognitive and motor impairments observed in affected individuals.

MRI offers a powerful, non-invasive approach to characterize brain morphology in rare genetic disorders. Advanced neuroimaging techniques, including volumetric analysis and cortical thickness measurements, can detect subtle structural alterations that may not be apparent on routine clinical imaging. In this study, real-world MRI scans were collected from 11 patients, with 16 scans sufficient for analysis after review and processing, for assessment of the natural history and progression of brain morphology in NGLY1 Deficiency. Patient ages ranged from 2 to 19 years of age at the time of the MRI. Five of the patients were female and six were male. Given the central role of NGLY1 in cellular protein homeostasis and the neurodevelopmental phenotype associated with its deficiency, we hypothesized that affected individuals would demonstrate distinct patterns of regional brain volume reductions and altered cortical morphometry compared to typically developing controls.

## Methods

### Participants

This was a real-world study in which the collected imaging was taken as part of the care of NGLY1 Deficiency patients outside of clinical trials. Participants were consented to share MRI under one of two protocols (Investigating the Biology and Natural History of the NGLY1 Related Disorder, #41906, or NGLY1 Deficiency: A Prospective Natural History Study, NCT03834987). The study protocol was approved by the Stanford Institutional Review Board (IRB Protocol # 47335). Parents gave written informed consent for their children or dependents. Enrollment criteria included a molecular diagnosis of NGLY1 Deficiency (either through genetic testing or a combination of genetic testing and biomarker confirmation). Imaging was requested and transferred through secure transfer or by disc.

### MRI collection

Participants were scanned at multiple sites around the world in the course of their medical care and there was no consistent protocol used to collect data. In total, 41 scans from 16 patients were assessed for potential inclusion. 28 were low resolution or had poor contrast or a cropped field of view. Three of these were rescued using SynthSeg, a tool from FreeSurfer to enhance the resolution of clinical-grade scans and generate more accurate segmentations.^15,16^ Scan details for the final 16 acceptable for inclusion are provided in **Supplementary Table 1**, including scan manufacturer and model, field strength, and voxel size. Half of the scans came from a 1.5T scanner and half were from a 3T scanner. Voxel resolution varied considerably, from 0.5 mm isotropic to inplane resolution=0.9×0.9 mm and slice thickness=6.5 mm.

### MRI processing

All MRI scans from patients over 3 years old were processed through FreeSurfer version 7.4.^17^ Most participants had multiple T1-weighted MRIs. All were processed and thoroughly visually quality checked. From the FreeSurfer aseg and aparc files we extracted: subcortical volume (7 regions bilateral), cortical surface area (31 regions bilateral), and cortical thickness (31 regions bilateral). The entorhinal cortex and parahippocampal gyrus were excluded from analyses as the medial surfaces of the temporal lobe are frequently poorly segmented. FreeSurfer is not recommended for participants under 4 years old, so the infant-specific version, infantFS, was used for younger patients.^18^ Image resolution varied, with multiple patients having voxel resolutions too large for good quality FreeSurfer segmentations; in these cases, SynthSeg was used to rescue these scans.^15,16^ SynthSeg is not available for infants, so analyses on patients <3 years old were completed only on native resolution scans. These results are reported in the supplement.

### Normative Modeling (patients >3 years old)

As there were no comparison participants scanned at each clinical site, we processed scans through CentileBrain, which compares values to a model trained on over 40,000 individuals between the ages of 3-90 years.^19–21^ This yielded Z-scores for each brain region, taking into account patient age and sex. Z-scores were calculated for cortical thickness, surface area, and subcortical volume.

### Normative Modeling (patients <3 years old)

The normative models in CentileBrain are not valid for individuals under 3 years old, and there are no publicly available tools that provide a similar function; therefore, we created our own normative models using data accessible through the National Institute of Mental Health (NIMH) Data Archive, or NDA. T1w MRIs were downloaded for all NDA participants between 6-36 months old for a total of 2,539 files. These were filtered to exclude scans from clinical samples (e.g. autism spectrum disorders) and others that did not fit the parameters, resulting in 573 MRIs that were processed through infantFS. InfantFS failed for 25 of 573 scans. This output was thoroughly visually quality checked, leading to 131 scans excluded for overall poor quality segmentation. For the remaining 417 scans, we noted any regional failures, using processes outlined by the ENIGMA Consortium.^22–24^ The most common areas of failure were the medial temporal lobes (as noted above) and the medial orbitofrontal cortex. For the scans for 6 and 7 month olds, the quality of the temporal lobe segmentation was poor in too many scans and therefore excluded. The number of control scans in the final model is shown in **Supplementary Table 2**, broken down by age and sex. As above, we extracted subcortical volume, cortical thickness, and cortical surface area from these scans, with the addition of cortical volume and curvature. For each sex, mean trajectories were modeled in R4.4.1 using basis splines with 3 degrees of freedom to capture developmental patterns while preventing overfitting. Standard deviation was estimated by fitting a separate spline model (2 degrees of freedom) to the absolute residuals from the mean model, with correction for the expected value of absolute deviations. This approach generated smooth, age-specific percentile curves (±1SD, ±2SD) that account for both mean changes and variance heterogeneity across development. Growth curves for each morphological measure as shown in **Supplementary Figures 1-7**, with separate curves for males and females. Trajectories for temporal lobe regions start at 11 months due to the sparsity of high quality segmentations as noted above. NGLY1 patient metrics were converted to standardized Z-scores by comparing observed values to the sex- and age-specific normative mean and standard deviation derived from these models. Z-scores were calculated for cortical thickness, surface area, curvature, and volume, and subcortical volume. Cortical curvature and volume were not available through CentileBrain for the older patient group.

### Additional variables

Patient birthweight and gestational age were collected, with birthweight divided into very low (<1.5kg), low (1.5kg-2.5kg), and normal (>2.5kg) categories. We additionally collected multiple phenotypes coded in human phenotype ontology (HPO) terminology. We examined the following HPO phenotypes: dysphagia (difficulty swallowing, HP:0002015), EEG abnormality (HP:0002353), gait disturbance (HP:0001288), generalized hypotonia (HP:0001290), hearing abnormality (HP:0000364), hyperkinetic movements (HP:0002487), seizure (HP:0001250), sleep disturbance (HP:0002360), and tremor (HP:0001337). Other HPO phenotypes were collected but not examined due to either no or limited variability across the samples (e.g. global developmental delay [HP:0001263] was present in all patients) or limited relevance to neuroimaging phenotypes (e.g. liver cirrhosis [HP:0001394]). Given the issues with data quality and the known motor phenotypes of NGLY1 Deficiency, we considered whether the patients whose MRI data was of sufficient quality were less severely impacted than the general NGLY1 Deficiency patient population. Comparing the incidence of each of these clinical variables between the sample with good quality MRI (N=11) vs. the full sample (N=16), we did not see significant deviations in the expected counts (*p*=.995). This supports the assumption that the patients included in the following analyses are generally reflective of the larger patient population.

### Statistical Analyses

We used a modified Bonferroni correction for multiple comparisons created by Li and Ji,^25^ which yielded the effective number of independent variables (V_eff_) in our analysis as 12 and a significance threshold of p < 0.05/12 = .0042. A traditional Bonferroni correction was too conservative for our analysis because there are correlations between test statistics (i.e., between the 62 regions examined). Our primary analysis was to examine whether cortical and subcortical morphology differed between the NGLY1 Deficiency patients and expected values. We conducted 2 sided *T*-tests on the *Z*-scores, testing whether they differed significantly from 0.

As secondary analyses we examined correlations between brain morphometry and the clinical variables listed above. Given the small sample size, binary nature of clinical variables, and exploratory nature of these analyses, Spearman rank correlations were computed to assess associations between clinical features and regional brain measures as Spearman correlation is more robust to outliers and does not assume normality of the data.

A graphic explaining the cortical metrics examined is shown in **Supplementary Figure 8**.

### Data Availability

The authors will share the data with qualified investigators whose proposal of data use has been approved by an independent review committee.

## Results

All analyses were performed separately for the scans from older (> 3 years old) versus younger (< 3 years old) patients, with some longitudinal patients having scans included in both phases.

### Primary Results

*Z*-scores are shown for subcortical volume, cortical thickness, and cortical surface area (**Figure 1**) and displayed projected onto the brain in **Figure 2**. Of the 10 scans included in the older cohort, 4 were enhanced with SynthSeg. We re-ran all analyses in the older cohort excluding these scans (**Supplementary Figures 9-12**).

**Figure 1.**
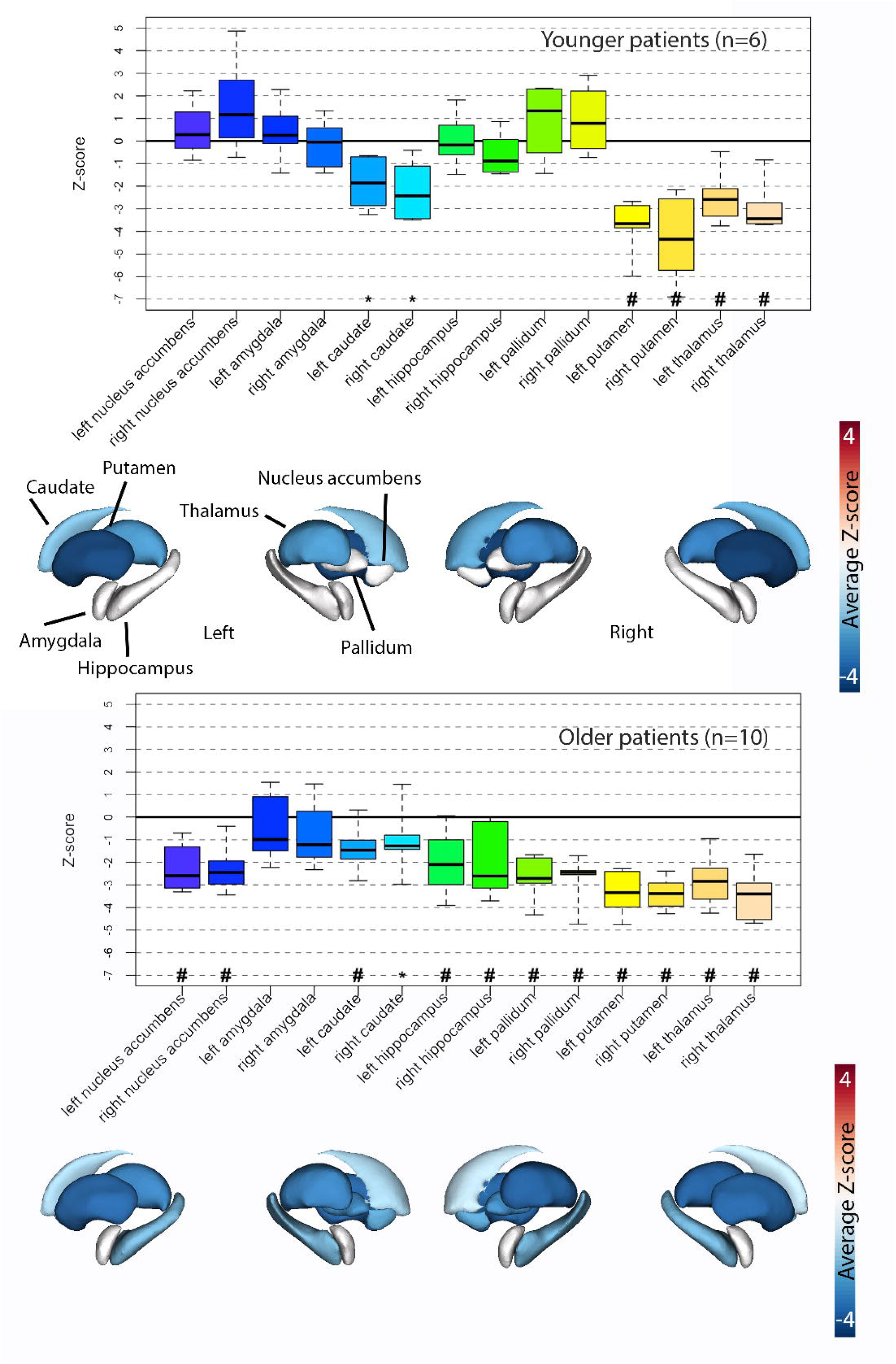
Subcortical volume Z-scores in patients. Boxplots display Z-scores for each subcortical structure (n=6 scans for 6 patients under 3 years old and 10 scans for 7 patients over 3 years old) relative to age- and sex-matched normative data. Box elements represent median, interquartile range (IQR), and whiskers extend to minimum and maximum excluding outliers; points beyond whiskers are outliers. Symbols indicate significant deviations from normative means: ^#^p<.05 after multiple comparisons correction (Li & Ji); *p<.05 uncorrected.

**Figure 2.**
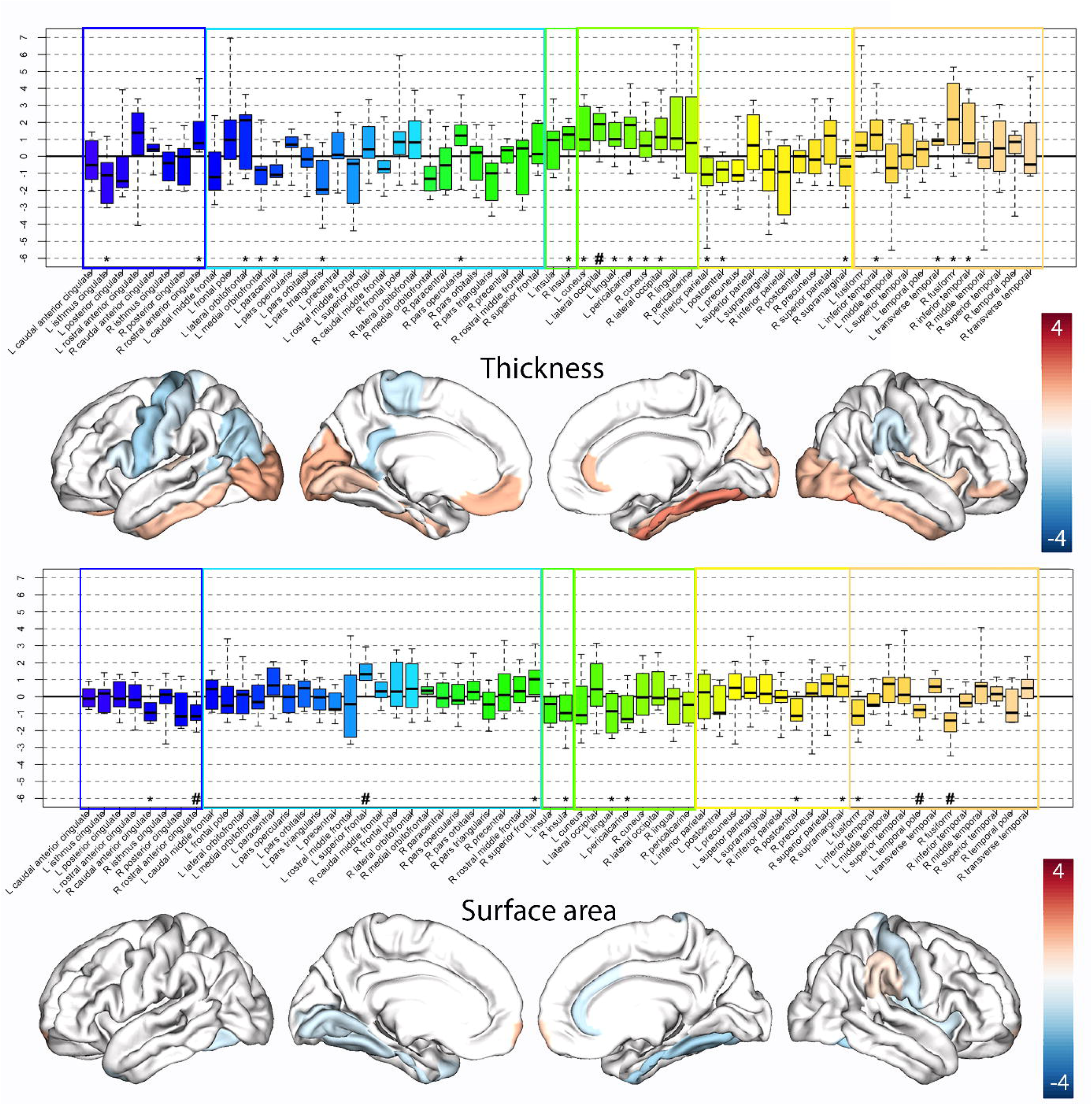
Cortical thickness and surface area effects for older patients. The average *Z-score*s for patients over 3 years old are displayed both in boxplot and projected onto the cortex for cortical thickness and surface area. Box elements (left panel) represent median, interquartile range (IQR), and whiskers extend to minimum and maximum excluding outliers; points beyond whiskers are outliers. Symbols indicate significant deviations from normative means: ^#^p<.05 after multiple comparisons correction (Li & Ji); *p<.05 uncorrected. Colored boxes indicate lobe - dark blue=cingulate, light blue=frontal, middle two=insula, green=occipital, yellow=parietal, dark yellow=temporal. For the cortical projections (right panel), marginally significant regions (.0042<*p*<.05) are included to display trends. Color corresponds to the average Z-score across the group for a given region, with negative Z-scores in blue and positive Z-scores in red. Left in image is left in brain.

In the younger patients, we found significantly smaller volumes in the bilateral putamen and thalamus (*t*-statistic=-5.2 -, −6.6, *p*<.0042), along with marginally lower volume in the bilateral caudate (*t=*-3.8 - −4.1, .0042<*p*<.05, **Figure 1**). We also found significantly lower volume bilaterally in older NGLY1 Deficiency patients in the accumbens, hippocampus, pallidum, putamen, and thalamus, and left caudate (*t*=-4.6 - −18.5), along with marginally lower volume in the right caudate (*t*=-3.2). Results in the subset of native resolution scans from the older patient group (n=7) were generally consistent, although differences in the left amygdala, left putamen, and right pallidum were no longer significant (**Supplementary Figure 9**).

For cortical thickness, in younger patients we found significantly thicker cortex in the right rostral middle frontal gyrus (*t*=8.5), and marginally thicker cortex in the left *pars opercularis, pars orbitalis*, and *pars triangularis*, along with the left cuneus, left superior parietal gyrus, and left supramarginal gyrus (*t*=3.1 - 8.5, **Figure 1**). We additionally found marginally thinner cortex in the bilateral isthmus of the cingulate (*t*=-2.7 - −3.1). In older patients, there was significantly thicker cortex in the left lateral occipital gyrus (*t*=4.7), along with marginally thicker cortex in the left rostral anterior cingulate, lateral orbitofrontal gyrus, and the right *pars opercularis*, insula, and multiple regions of the occipital and temporal cortex (*t*=2.3-3.5). In addition, we found marginally thinner cortex in the left isthmus of the cingulate, medial orbitofrontal gyrus, paracentral gyrus*, pars triangularis*, inferior parietal gyrus, and postcentral gyrus, and the right supramarginal gyrus (*t*=-2.3 - −3.3). For the older patients, cortical thickness effects appear to cluster as thinner cortex in dorsal brain regions and thicker cortex in ventral brain regions. Results are largely consistent when limiting analyses in the older cohort to native resolution scans (**Supplementary Figure 10**).

For surface area, we saw widespread areas of significantly lower surface area in the younger patients, including multiple sections of the cingulate gyrus, the bilateral insula, and multiple gyri within the frontal and parietal cortices (*t*=-5.1 - −13.6), with additional areas of marginally lower surface area (*t*=-2.6 - −5.0). Differences in the older patients were less pronounced, with significantly lower surface area in the left temporal pole and right rostral anterior cingulate and fusiform gyrus (*t*=-3.9 - −4.9), and marginally lower surface area in the left lingual, pericalcarine, and fusiform gyri, and the right caudal anterior cingulate, insula, postcentral gyrus (*t*=-2.3 - −3.8). Surface area was significantly higher in the left superior frontal gyrus (*t*=5.6) and marginally higher in the right superior frontal gyrus, along with the right supramarginal gyrus (*t*=3.0 - 3.2). Results were consistent in the native resolution only subset (**Supplementary Figure 11**). All of the results comparing the native resolution subset to the full older patient sample are displayed in **Supplementary Figure 12**.

Given the widespread differences in the younger patient cohort, we examined two additional cortical morphology measures: curvature and volume. Consistent with the effects for cortical thickness and surface area, we found significantly lower cortical curvature and volume in the NGLY1 Deficiency patients compared to typically developing infants and babies (complete results in **Figure 3**). There was significantly lower curvature in the right *pars triangularis*, rostral middle frontal, superior frontal, and supramarginal gyrus, and the left insula (*t*=-2.6 - −4.4), along with marginally lower curvature across regions of the cingulate cortex, left frontal cortex, right insula, lateral occipital gyrus, left supramarginal gyrus, bilateral inferior temporal gyrus, and right fusiform gyrus (*t*=-5.1 - −16.9). Volume was also significantly lower in the bilateral precentral gyrus, insula, precuneus, right cingulate cortex, fusiform gyrus, paracentral gyrus, *pars opercularis* (*t*=-5.1 - −9.2), and was marginally lower in the bilateral superior frontal gyrus, left caudal middle frontal, paracentral, *pars opercularis*, cingulate cortex, lateral occipital, superior parietal, fusiform, middle temporal gyrus, and right inferior parietal gyrus (*t*=-2.6 - −4.8).

**Figure 3.**
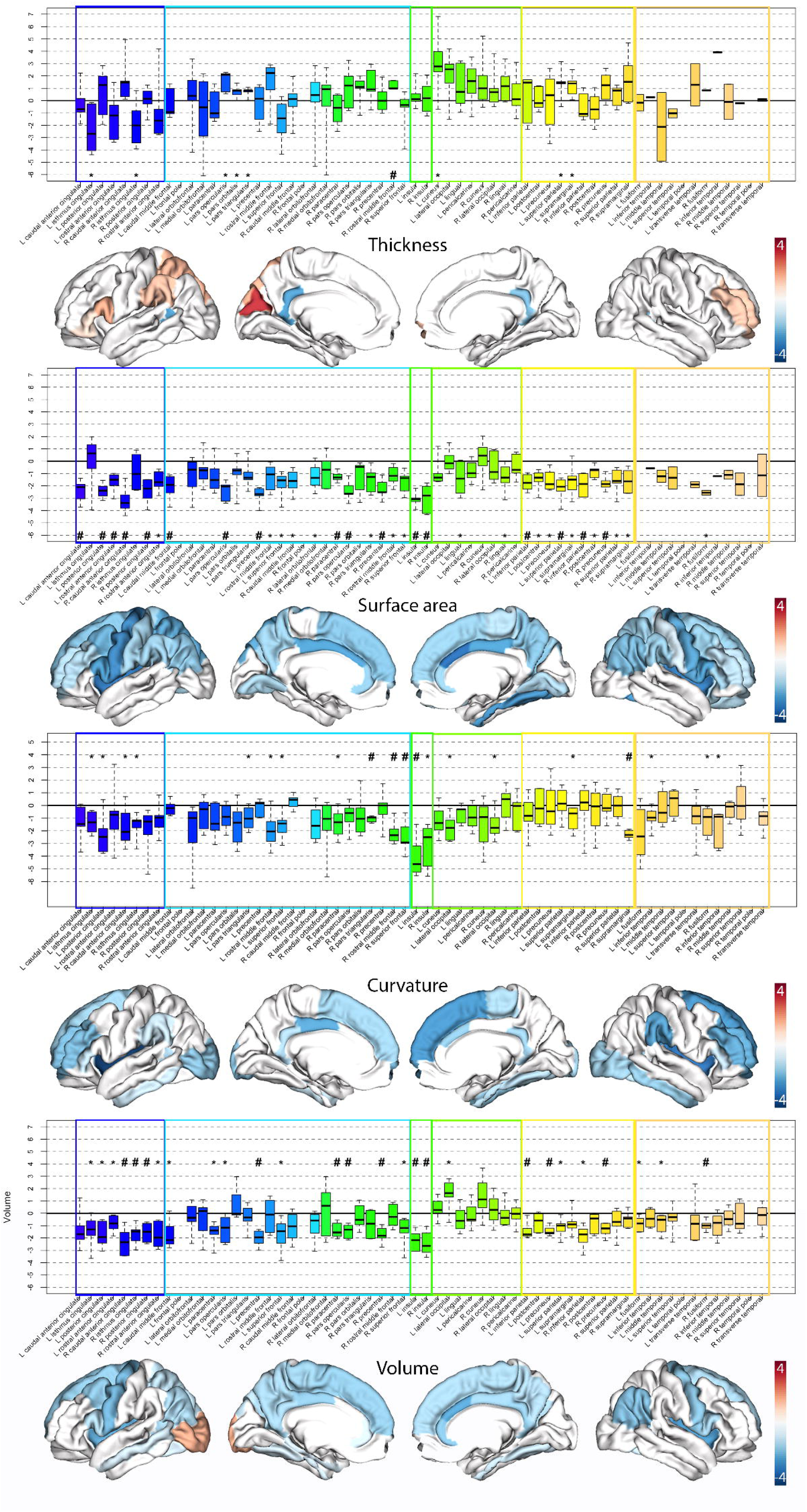
Complete cortical morphology effects for younger patients. The average Z-scores for patients under 3 years old are displayed both in boxplot and projected onto the cortex for cortical thickness, surface area, curvature, and volume. Box elements (left panel) represent median, interquartile range (IQR), and whiskers extend to minimum and maximum excluding outliers; points beyond whiskers are outliers. Symbols indicate significant deviations from normative means: ^#^p<.05 after multiple comparisons correction (Li & Ji); *p<.05 uncorrected. Colored boxes indicate lobe - dark blue=cingulate, light blue=frontal, middle two=insula, green=occipital, yellow=parietal, dark yellow=temporal. For the cortical projections (right panel), marginally significant regions (.0042<*p*<.05) are included to display trends. Color corresponds to the average Z-score across the group for a given region, with negative Z-scores in blue and positive Z-scores in red. Left in image is left in brain.

### Clinical Correlations

Given the limitations of the SynthSeg data as mentioned above, we examined associations between cortical and subcortical morphology and clinical measures using the native resolution data only. Separate analyses were run for younger and older patients. In order to limit the number of statistical comparisons and to focus on clinically-relevant associations, we limited the analysis to those regions that had a *p*-values below 0.10 in the primary results above. The top 10 associations for younger and older patients are reported in **Figure 4**, and the rest of the results in **Supplementary Table 3** and **4**.

**Figure 4.**
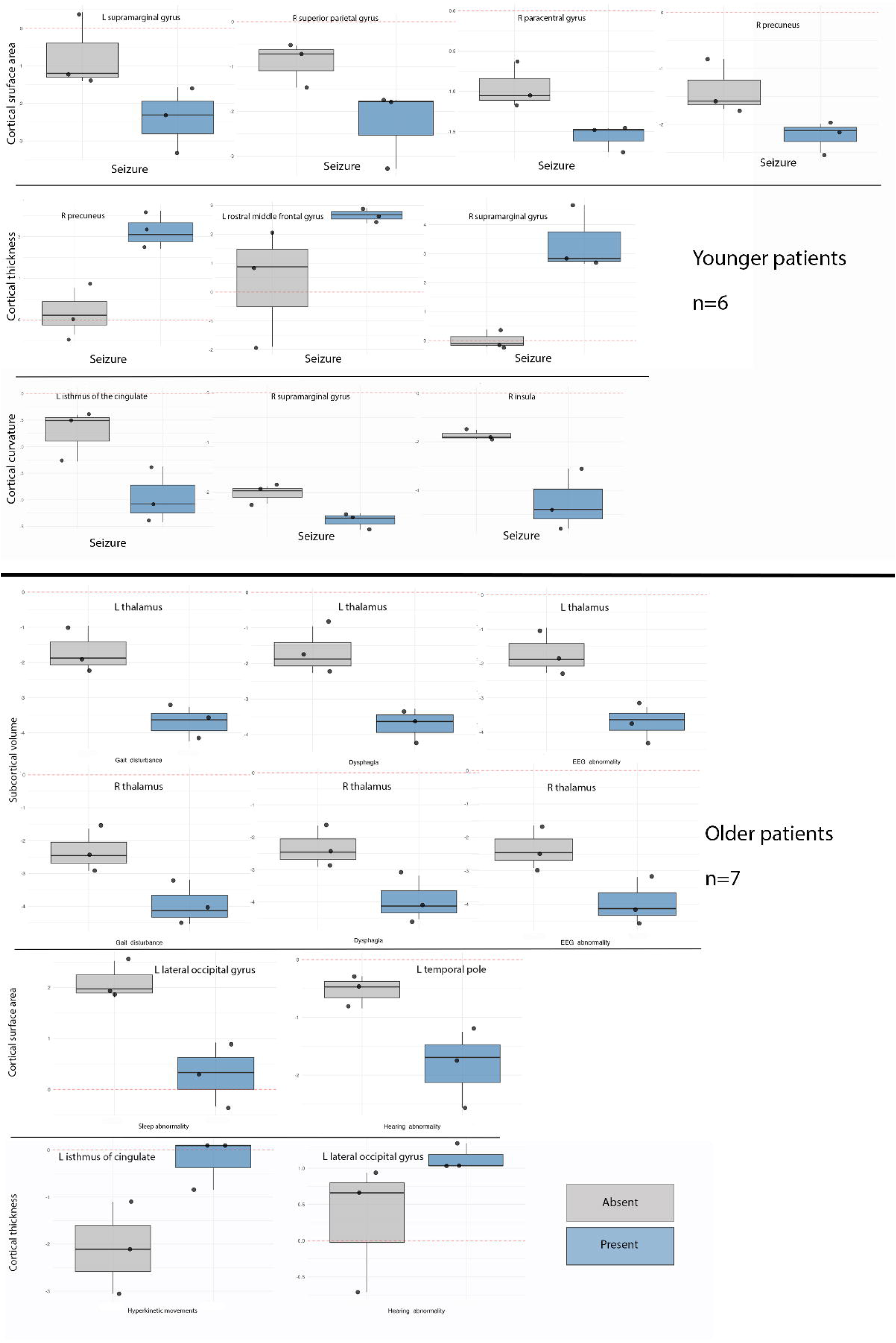
Clinical associations. Associations between cortical surface area (top), cortical thickness (middle), and cortical curvature (bottom) with clinical variables in the younger (top) and older (bottom) patient group. Boxplots demonstrate differences in Z-score based on the presence (blue) vs. absence (gray) of seizure and gait disturbance.

Several brain regions showed identical correlation coefficients and p-values with various clinical variables. This reflects the rank-based nature of Spearman correlation: although the absolute Z-score values differed across regions, the relative ranking of subjects remained consistent, resulting in identical test statistics. In the younger patients, seizure was associated with thicker cortex in the right precuneus, right supramarginal gyrus, and left rostral middle frontal gyrus (⍰s=.88, *p*s<.021), along with lower surface area in the left supramarginal, right paracentral, and right superior parietal gyri, and right precuneus (⍰s=-.88, *p*s<.021), and lower curvature in the left isthmus of the cingulate, right supramarginal gyrus, and right insula (⍰s=-.88, *p*s<.021). In the older patients, smaller volume for the bilateral thalamus was associated with several clinical variables - gait disturbance, dysphagia, and EEG abnormality (⍰s=-.88, *p*s<.021). Seizure was associated with greater surface area in the right insula (⍰=.88, *p*<.021). Lastly, hearing abnormality was associated with smaller surface area in the left temporal pole (⍰=-.88, *p*<.021), thicker cortex in the left transverse temporal gyrus (⍰=.88, *p*<.021), and thinner cortex in the left lateral occipital gyrus (⍰=-.88, *p*<.021).

## Discussion

Here we describe alterations in the cortical and subcortical morphology of patients with NGLY1 Deficiency, an ultra-rare disorder that results in global developmental delay and is associated with a significantly shortened life expectancy (median age at death=13 years old).^3^ This comprehensive examination of brain structural abnormalities in this patient population to date reveals pervasive alterations in cortical and subcortical development. The most consistent finding across age ranges was smaller subcortical volumes, particularly for the thalamus but also including the caudate and putamen. This change in the thalamus may have a wide range of clinical consequences, given the broad functions (e.g., cognition, motor, sensory processing) and connectivity of the thalamus, as smaller thalamic volume in the older patients was associated with a number of clinical features of NGLY1 Deficiency.

Effects in the cortex varied with age, with different patterns emerging in the younger vs. older patient groups. Prior MRI studies in NGLY1 Deficiency have largely been case reports. Results from these have varied, with some having minimal if any structural abnormalities,^8,9^ while others have noted mild atrophy^6,9–11^ or thin spinal cord.^12^ One case report described more widespread atrophy along with white matter abnormalities including prominent perivascular spaces,^13^ while a relative expansion of fluid-filled spaces both in and around the brain.^14^

In the younger patients, there was a trend towards thinner cortex in the cingulate and scattered areas of thicker cortex, including the right inferior frontal gyrus (*pars opercularis, orbitalis,* and *triangularis*) and left parietal cortex. There were widespread areas of lower surface area in the younger patients, encompassing broad areas of the frontal, parietal, cingulate, and insular cortex, with minimal effects in the temporal cortex and relative sparing of the occipital cortex. Additionally, cortical curvature and volume were lower across large areas of cortex. Lower curvature indicates less cortical gyrification, with wider, flatter gyri and shallower sulci. Taken together, these results fit with prior studies indicating that NGLY1 Deficiency is associated with significant underdevelopment in brain structure.

Prior studies using patient stem cell-derived organoids showed a reduction in neuronal and astrocytic protein markers,^26^ suggesting that NGLY1 Deficiency may impair late-stage neurogenesis and the subsequent migration of neurons to the upper cortical layers.^27^ Impairments in neurogenesis, neuronal differentiation, and neuronal survival would have broad implications for brain structural development. Neuronal migration and layering is dependent on the guidance of radial glial cells,^28,29^ which also serve as progenitors for multiple neuronal populations, but the signaling necessary to maintain these are disrupted in cerebral organoids developed from NGLY1-deficient stem cells.^27^ Differential expansion of the inner versus outer cortical layers contributes to cortical folding,^28^ but upper-layer neurons are affected in these organoids.^27^ Synaptic pruning is another key process shaping brain development, and it occurs at varying rates across the brain.^30,31^ Disruptions of protein degradation, proteostasis, and proteasome function that result from NGLY1 Deficiency could impair the synaptic pruning process.^26^ Taken together, there are multiple cellular mechanisms that are altered in NGLY1 Deficiency, and any or all of them could be responsible for the cortical and subcortical deficits we report.

In the older patients, there was a trend towards thinner cortex in dorsal brain regions such as the lateral parietal and occipital cortex, and thicker cortex in ventral regions such as the inferior temporal and occipital cortex. Examining gradients of cortical thickness over normal development, cortex is thickest in the dorsal frontal and temporal regions and is thinnest in the parietal and medial occipital cortex.^32^ Further, cortex thins fastest in the frontal pole and medial parietal cortex and thins slowest in the lateral parietal cortex during typical development. This could indicate an acceleration of the typical developmental pattern in affected patients. The cerebellum, which is one of the primary areas of interest in NGLY1 Deficiency given the loss of Purkinje cells, is highly connected to the cerebrum through the thalamus, the region where the most consistent deficits were observed.^33,34^ Specific thalamic nuclei project to distinct cortical targets, so the variable effects on cortical morphology could be influenced by differential degeneration of different thalamic nuclei. Unfortunately, we did not have the resolution to examine effects in this level of detail, but future analyses may be able to collect more specific data.

Interestingly, while younger patients showed widespread reductions in surface area across multiple brain regions compared to expected values, these differences were largely absent in the older patient cohort. This age-related divergence in cortical phenotypes raises important interpretive questions. With limited longitudinal data available, we cannot definitively determine whether these varying effects reflect delayed neurodevelopment that partially normalizes over time, or whether they result from survivor bias given the severe nature of this disorder. The latter interpretation is particularly relevant considering that the median lifespan of patients with NGLY1 Deficiency is 13 years, with three patients in our cohort dying between ages 14-22 years. Those who survive into adolescence and young adulthood may represent a phenotypically distinct subgroup with less severe cortical alterations, potentially explaining the more limited surface area reduction observed in the older cohort.

Both the younger and older patients displayed significantly lower subcortical volume than expected values, suggesting consistent subcortical underdevelopment in NGLY1 Deficiency. The subcortical regions that seemed to have the most robust effect sizes were the caudate, putamen, and thalamus, which support a wide range of functions including motion, cognition, and emotion. The subcortical volume reductions we observed, particularly in the thalamus, align with neuropathological findings reported in NGLY1 Deficiency. An autopsy case report of a 5-year-old with NGLY1 Deficiency demonstrated neuronal protein accumulations throughout subcortical brain regions and extensive cerebellar cell loss.^34^ These protein accumulations likely result from the impaired proteostasis that characterizes NGLY1 Deficiency. Prior studies in mice have found poor brain development and accelerated neuronal degeneration, particularly in the thalamus,^35–37^ while another recent study in rats reported evidence of widespread neuroinflammation, particularly in subcortical regions.^38^ The correlation between our volumetric findings and these previously reported pathological changes suggests that the reduced subcortical volumes we detected may reflect underlying neuronal loss and dysfunction secondary to protein accumulation.^34^

When focusing on regions showing trend level alterations in the group analysis (*p*<.10, **Supplementary Tables 3 and 4**), we found that thicker cortex, lower surface area, and lower curvature were associated with the presence of seizures in the younger cohort. Other seizure disorders have some overlapping cortical patterns. For example, focal cortical dysplasia is a congenital brain malformation that leads to focal cortical thickening and often presents with severe, medication-resistant seizures,^39^ and childhood absence seizures are also associated with thicker cortex and lower volumes.^40–42^ Treatment-resistant seizures are common in NGLY1 Deficiency, with only ⍰ of patients achieving full control of their seizures.^43^ In older patients, the most consistent finding was bilateral thalamic volume reduction associated with gait disturbance, dysphagia, and EEG abnormality. However, because these features co-occurred in the same patients, our small sample size prevents us from determining whether this association reflects overall disease severity or is specific to any one clinical variable.

### Limitations

This study has several important limitations. The small sample size (n=11) limits statistical power and generalizability, though it reflects the ultra-rare nature of NGLY1 Deficiency with ∼165 documented cases worldwide. Data acquisition challenges introduced potential variability. Participants were each scanned at a different site using varying scanner models and protocols, with some undergoing lower resolution clinical-quality scans rather than higher resolution research-quality scans. This heterogeneity in scan resolution limited our ability to correlate neuroimaging findings with clinical phenotypes. Age-related methodological constraints further complicated analyses. We employed different processing pipelines for younger (< 3 years old) and older participants (> 3 years old) due to developmental differences in brain morphology and tissue contrast. Our normative reference models were not continuous across age groups, limiting longitudinal interpretations of *Z-score*s. The normative model for younger participants was based on a smaller reference control sample (n=417) and exhibited poor segmentation quality in temporal lobes for 6- and 7-month-old infants, leading us to exclude all normative data for the temporal lobes for the youngest participants. Finally, cerebellar analyses were not performed due to poor image quality in this region across multiple participants. Given pronounced motor impairments in NGLY1 Deficiency, the cerebellum’s role in motor coordination, and prior studies showing loss of Purkinje cells in NGLY1 Deficiency,^44^ this represents an important gap that should be addressed in future studies with optimized cerebellar imaging protocols. Despite these limitations, this study provides valuable insights into the neuroanatomical correlates of this ultra-rare disorder. Future multi-site collaborative studies with standardized protocols, larger sample sizes, and comprehensive phenotyping will be essential to validate and extend these findings.

This comprehensive neuroimaging characterization of NGLY1 Deficiency reveals pervasive alterations in brain development that extend beyond the relatively nonspecific atrophy previously reported in case studies. These findings establish a neuroimaging phenotype characterized by consistent subcortical volume reductions across age groups, particularly affecting the thalamus, caudate, and putamen, with age-dependent patterns of cortical alterations including widespread surface area reductions and altered gyrification in younger patients. The correlations between cortical and subcortical morphological markers suggest that quantitative neuroimaging may serve as a valuable biomarker for disease progression and therapeutic monitoring in future clinical trials. While our sample size was limited by the ultra-rare nature of this disorder, and interpretation is complicated by potential survivor bias, these findings provide critical insights into the neurodevelopmental trajectory of NGLY1 Deficiency. The distinct patterns of brain morphological changes identified may reflect the disrupted cellular processes inherent to this disorder, including impaired protein degradation, altered neuronal migration, impaired neuronal differentiation or survival, and abnormal synaptic development. As potential therapies for NGLY1 Deficiency advance toward clinical trials, the neuroimaging metrics established here may prove essential for objectively measuring treatment response and understanding the relationship between brain structure and functional outcomes in this devastating pediatric disorder.

## Supporting information

STROBE

## Acknowledgements

Thank you to the Grace Science Foundation for funding. Thank you to the patients and their families for their trust and support.

## Supplementary Material

**Supplementary Figure 1.**
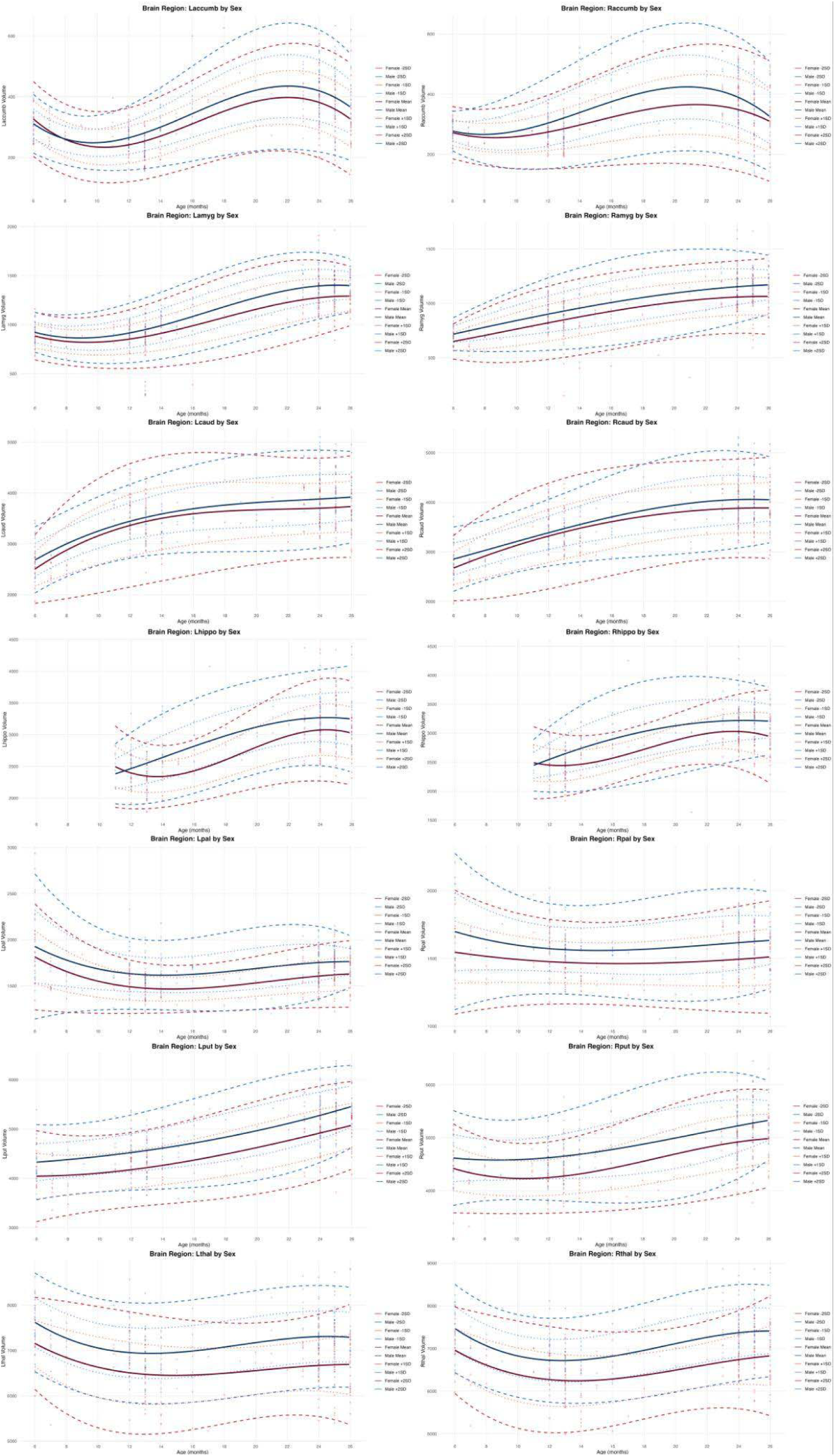
Normative trajectories of subcortical volume for ages 6-27 months. Normative trajectories created from 417 infants, separated by sex. Mean, 1 standard deviation (SD), and 2SD curves are shown for males (blue) and females (red).

**Supplementary Figure 2.**
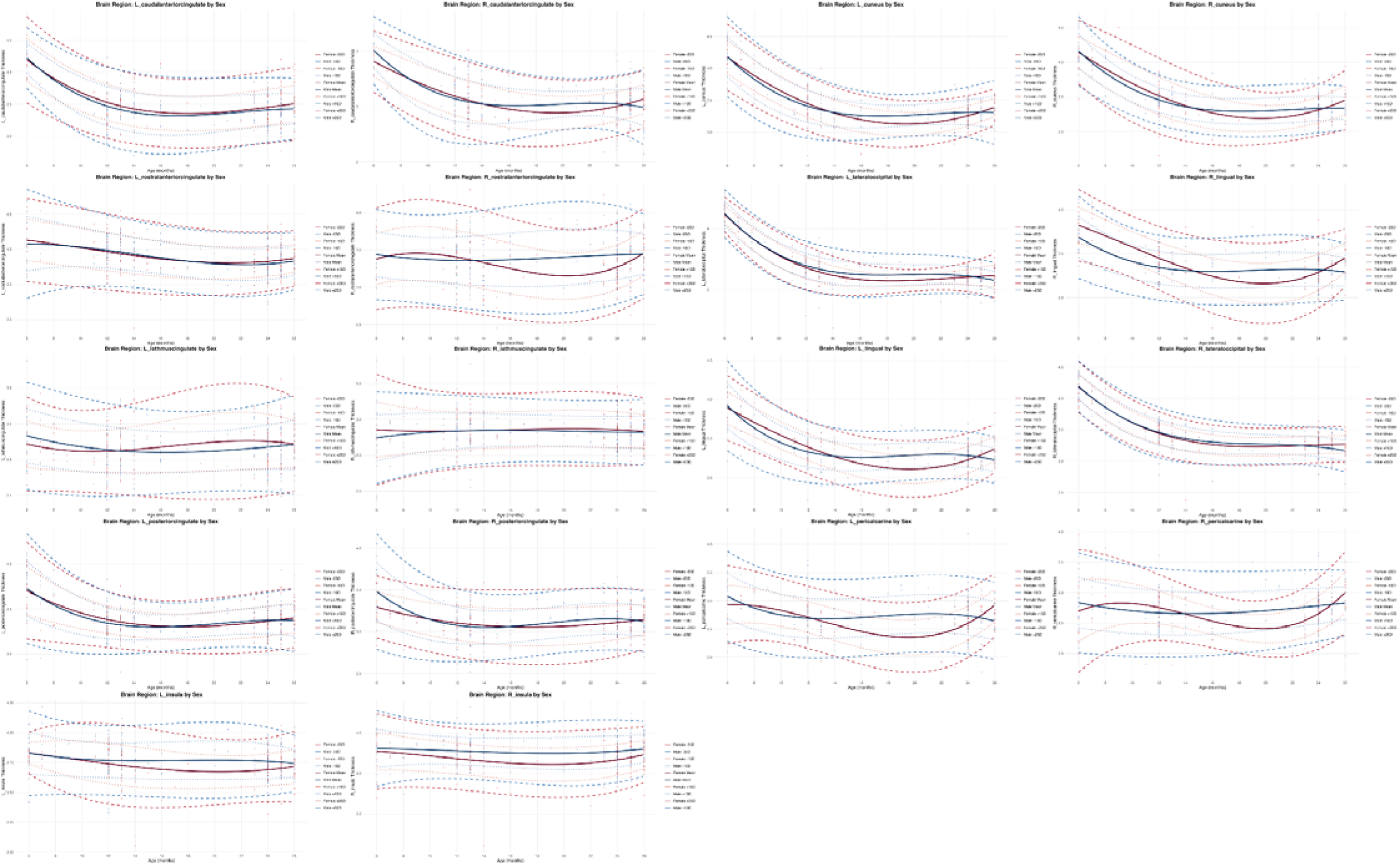
Normative trajectories of cortical thickness (cingulate, insula, and occipital lobes) for ages 6-27 months. Normative trajectories created from 417 infants, separated by sex. Mean, 1 standard deviation (SD), and 2SD curves are shown for males (blue) and females (red).

**Supplementary Figure 3.**
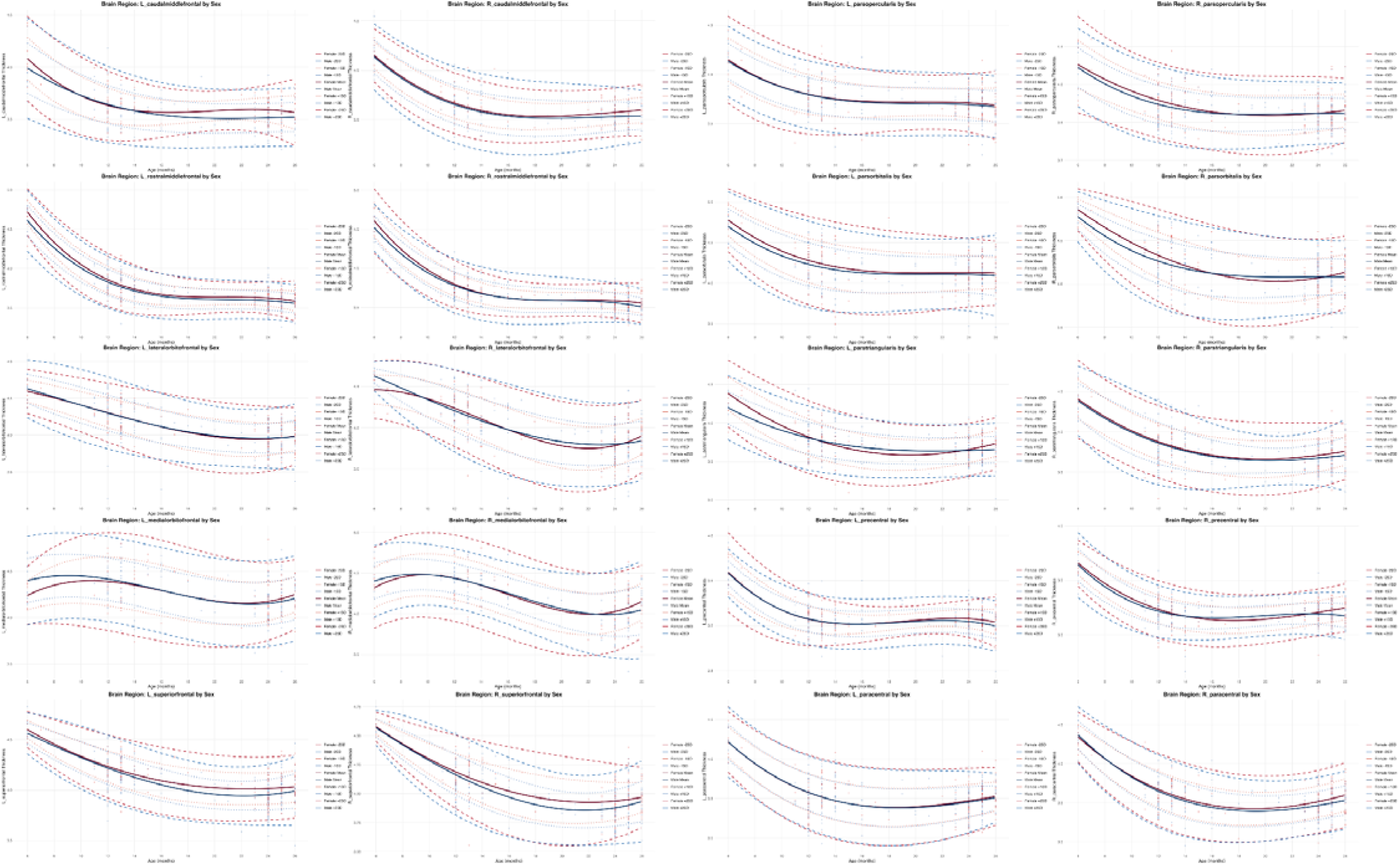
Normative trajectories of cortical thickness (frontal lobe) for ages 6-27 months. Normative trajectories created from 417 infants, separated by sex. Mean, 1 standard deviation (SD), and 2SD curves are shown for males (blue) and females (red).

**Supplementary Figure 4.**
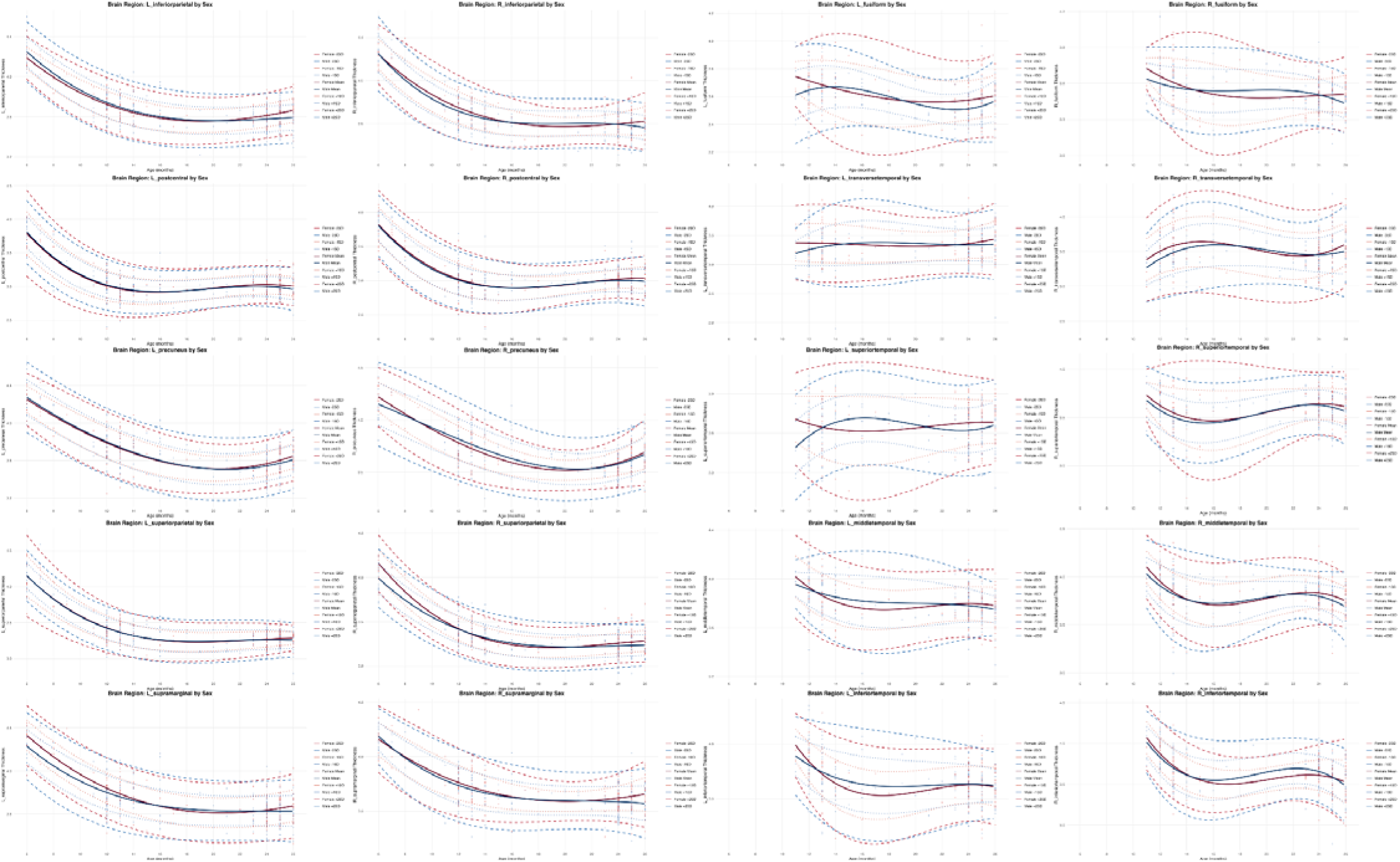
Normative trajectories of cortical thickness (parietal and temporal lobes) for ages 6-27 months. Normative trajectories created from 417 infants, separated by sex. Mean, 1 standard deviation (SD), and 2SD curves are shown for males (blue) and females (red).

**Supplementary Figure 5.**
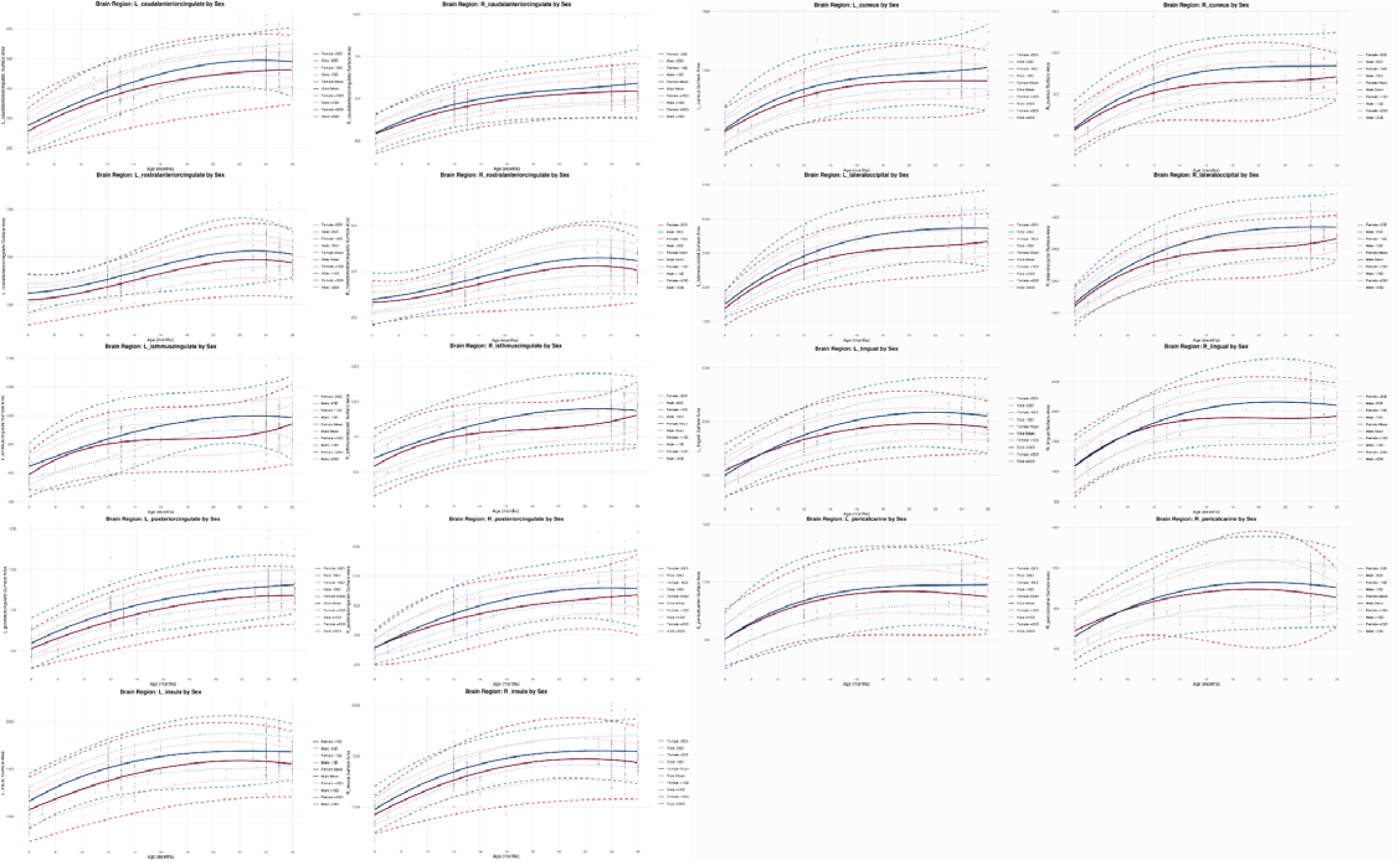
Normative trajectories of cortical surface area (cingulate, insula, and occipital lobes) for ages 6-27 months. Normative trajectories created from 417 infants, separated by sex. Mean, 1 standard deviation (SD), and 2SD curves are shown for males (blue) and females (red).

**Supplementary Figure 6.**
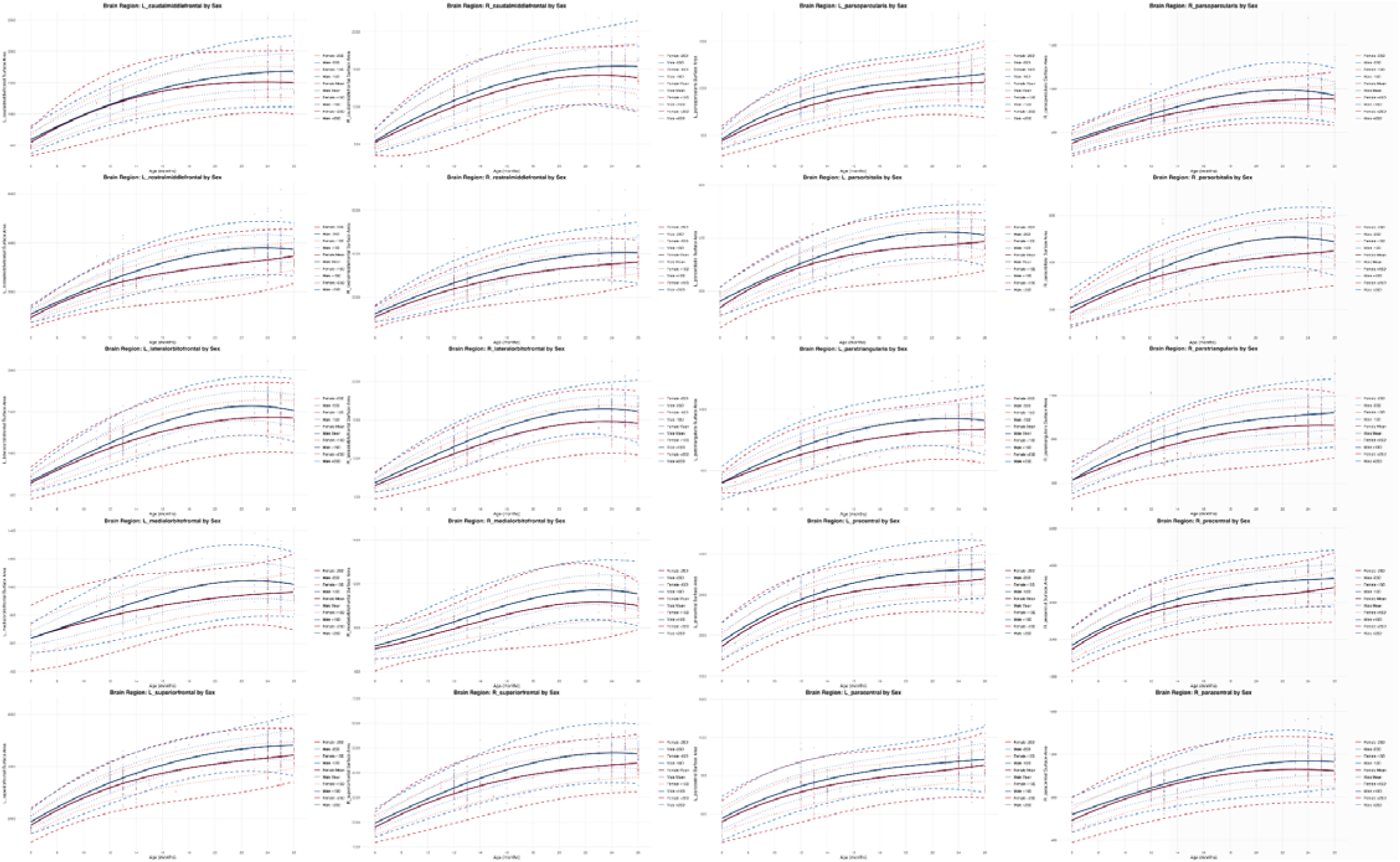
Normative trajectories of cortical surface area (frontal lobe) for ages 6-27 months. Normative trajectories created from 417 infants, separated by sex. Mean, 1 standard deviation (SD), and 2SD curves are shown for males (blue) and females (red).

**Supplementary Figure 7.**
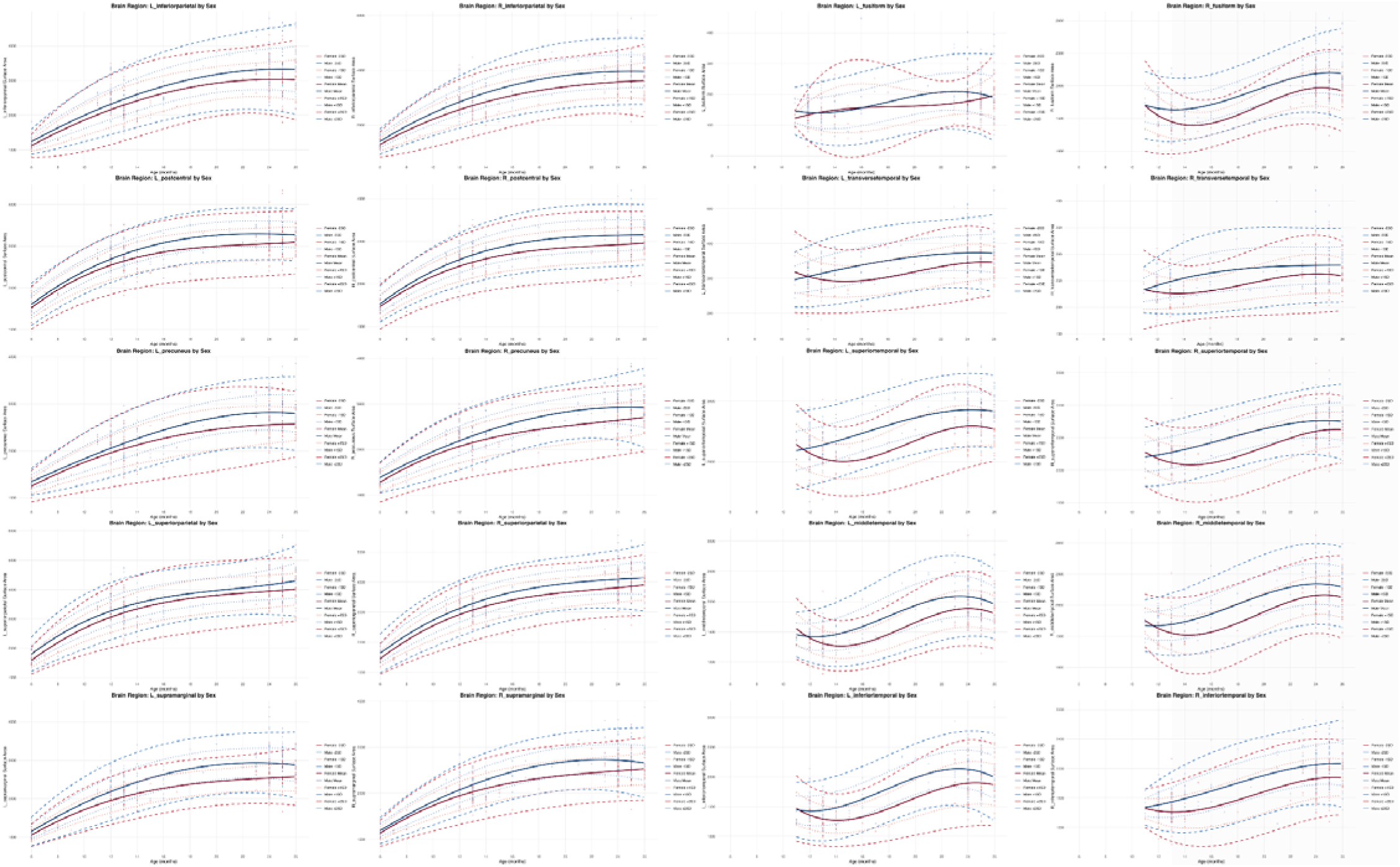
Normative trajectories of cortical surface area (parietal and temporal lobes) for ages 6-27 months. Normative trajectories created from 417 infants, separated by sex. Mean, 1 standard deviation (SD), and 2SD curves are shown for males (blue) and females (red).

**Supplementary Figure 8.**
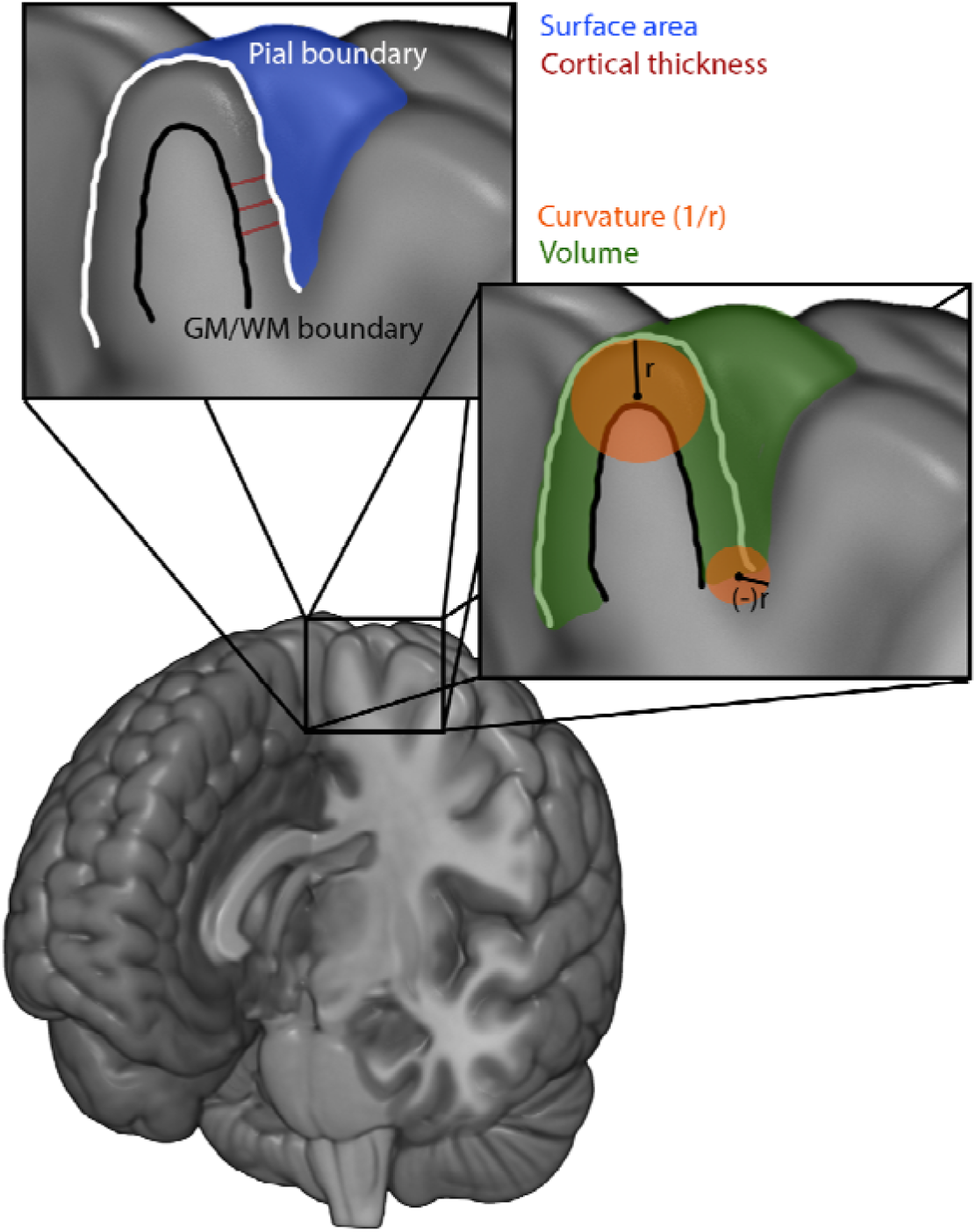
Visualization of the cortical morphometry metrics examined in this paper. The pial boundary for an individual gyrus is outlined in the coronal pop out in white, with the boundary between gray and white matter outlined in black. The distance between these, shown in the red lines, is the cortical thickness. The surface area is the area of the gray matter surface, as shown in blue. Volume is the gray matter volume of a region, as shown in green. Curvature is local folding of the cortex and is defined by the radius of the sphere that fits the curve. Mean curvature is the average of the first two principal components of curvature for a region, which are orthogonal to each other.

**Supplementary Figure 9.**
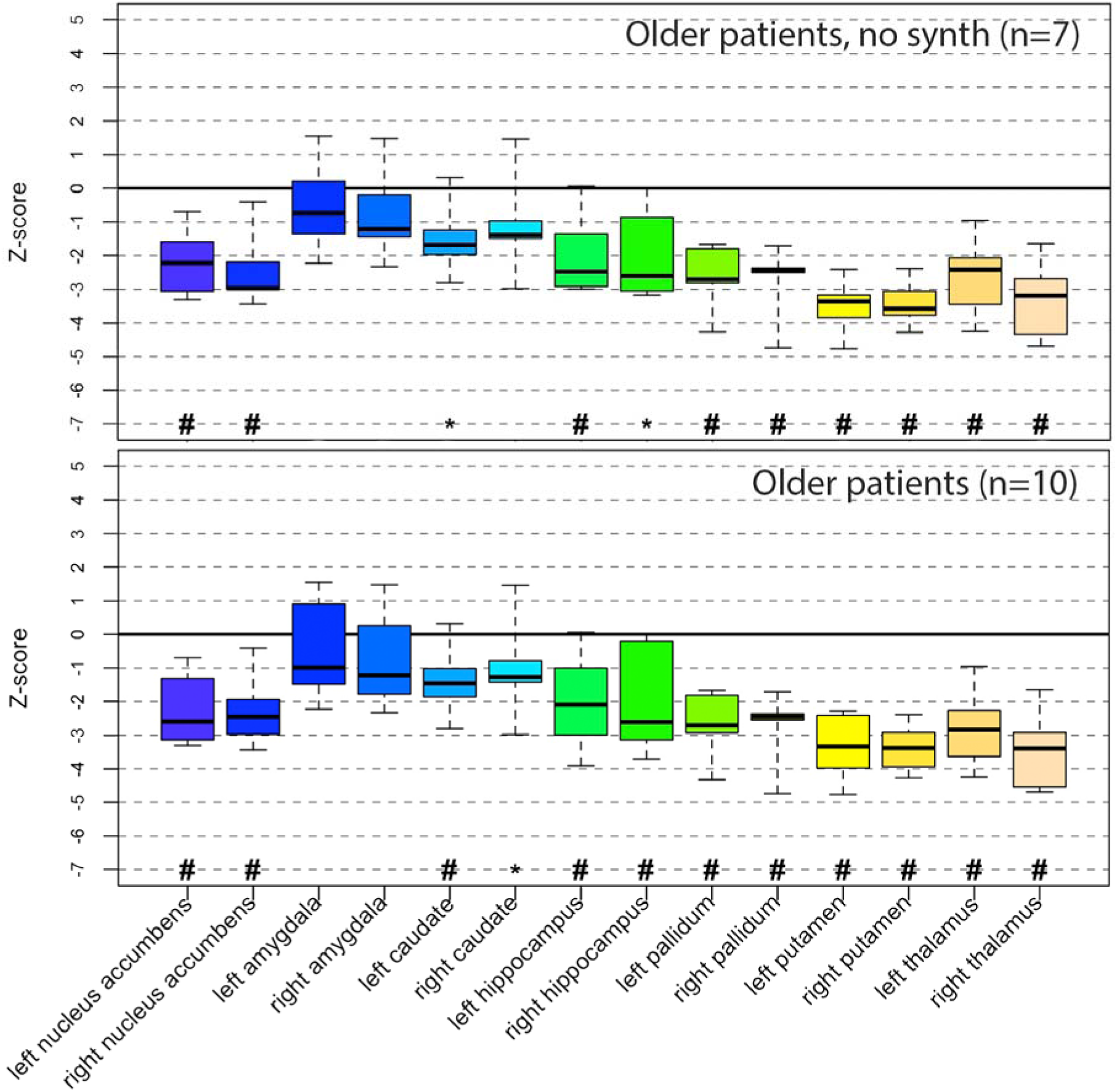
Subcortical volume Z-scores in older patients (real only and combined). Boxplots display Z-scores for each subcortical structure (top: only native resolution scans, bottom: native and SynthSR enhanced scans) relative to age- and sex-matched normative data. Box elements represent median, interquartile range (IQR), and whiskers extend to minimum and maximum excluding outliers; points beyond whiskers are outliers. Symbols indicate significant deviations from normative means: ^#^p<.05 after multiple comparisons correction (Li & Ji); *p<.05 uncorrected.

**Supplementary Figure 10.**
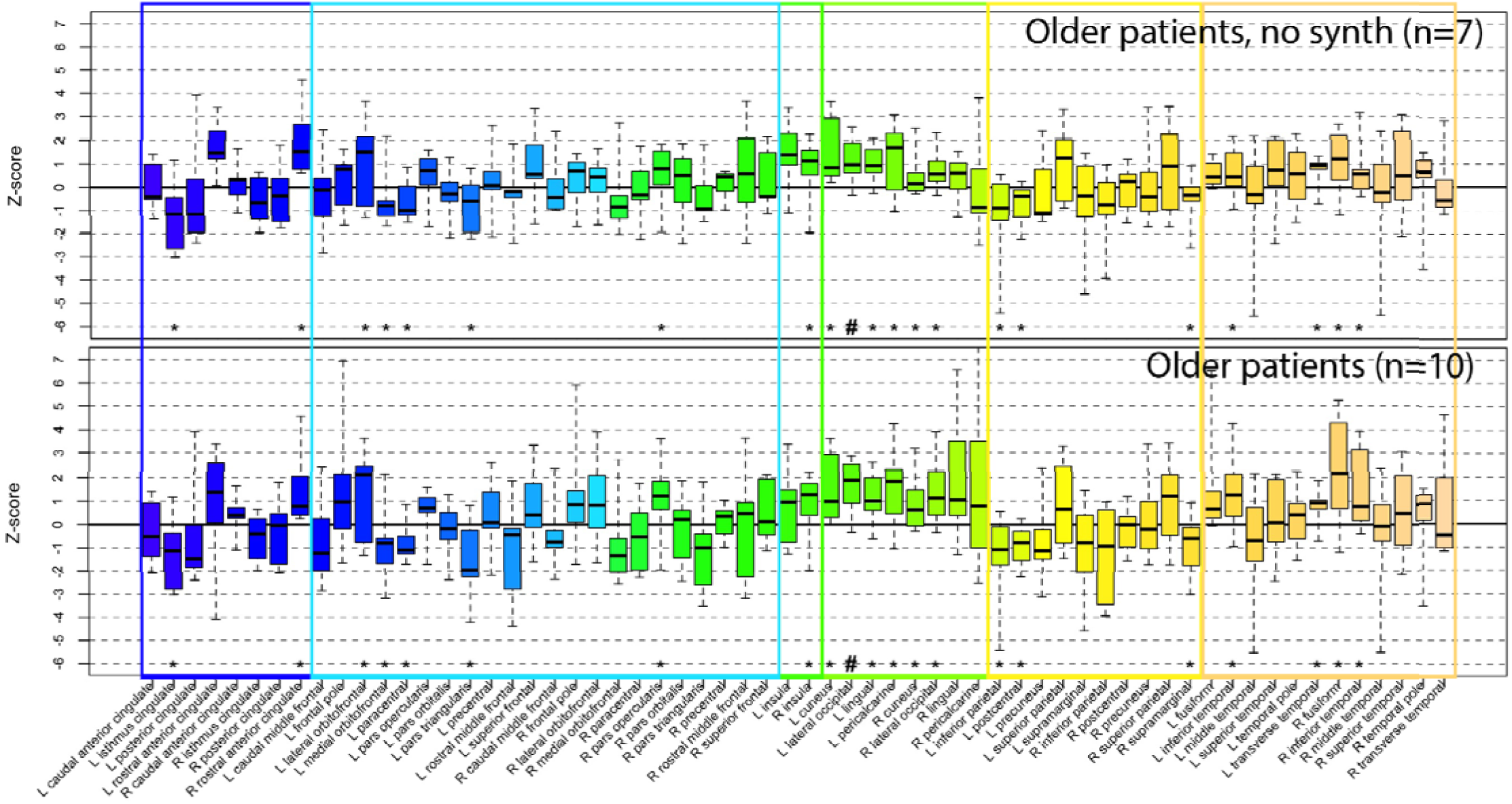
Cortical thickness Z-scores in older patients (real only and combined). Boxplots display Z-scores for each cortical region (top: only native resolution scans, bottom: native and SynthSR enhanced scans) relative to age- and sex-matched normative data. Box elements represent median, interquartile range (IQR), and whiskers extend to minimum and maximum excluding outliers; points beyond whiskers are outliers. Symbols indicate significant deviations from normative means: ^#^p<.05 after multiple comparisons correction (Li & Ji); *p<.05 uncorrected.

**Supplementary Figure 11.**
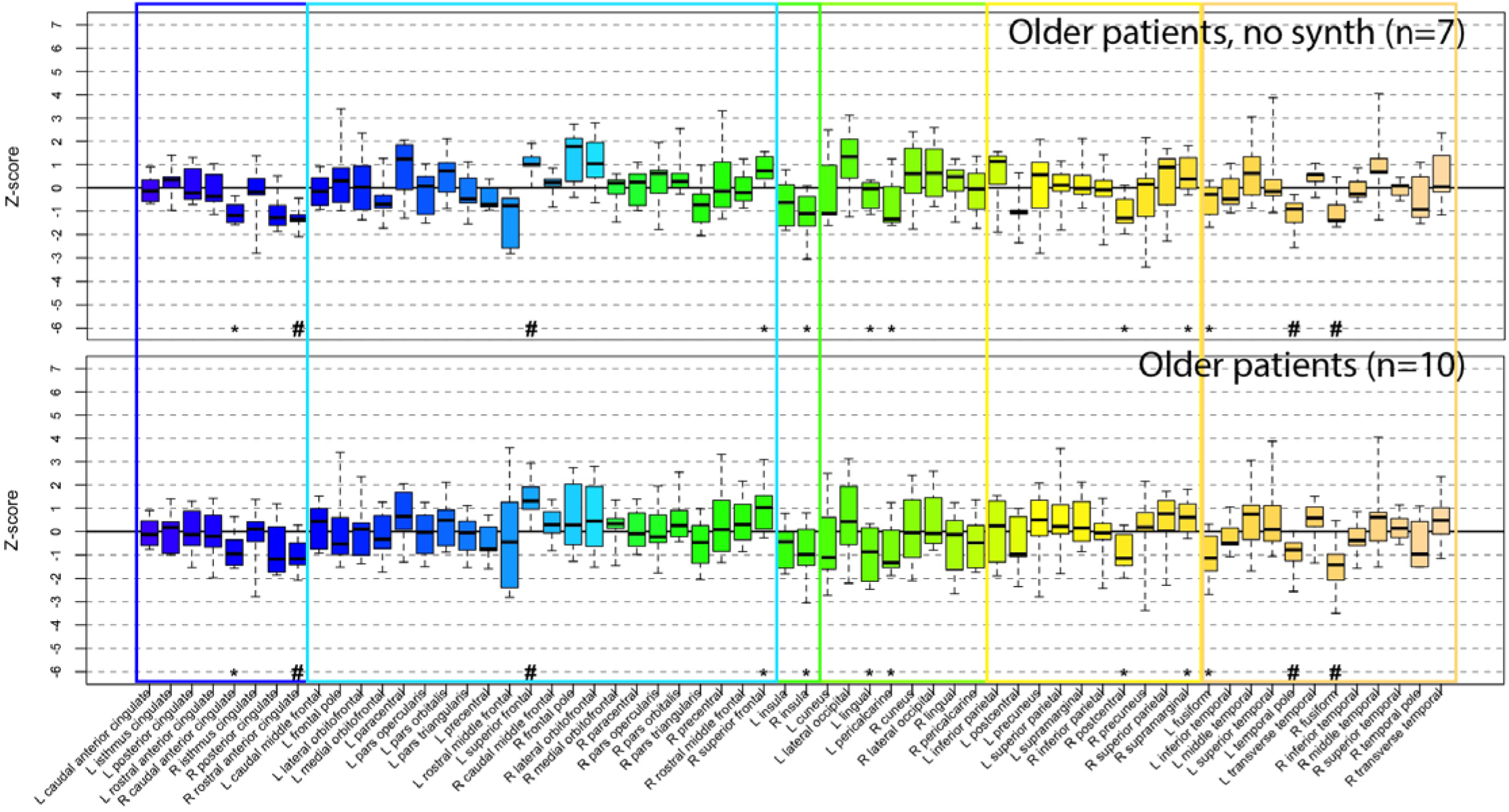
Cortical surface area Z-scores in older patients (real only and combined). Boxplots display Z-scores for each cortical region (top: only native resolution scans, bottom: native and SynthSR enhanced scans) relative to age- and sex-matched normative data. Box elements represent median, interquartile range (IQR), and whiskers extend to minimum and maximum excluding outliers; points beyond whiskers are outliers. Symbols indicate significant deviations from normative means: ^#^p<.05 after multiple comparisons correction (Li & Ji); *p<.05 uncorrected.

**Supplementary Figure 12.**
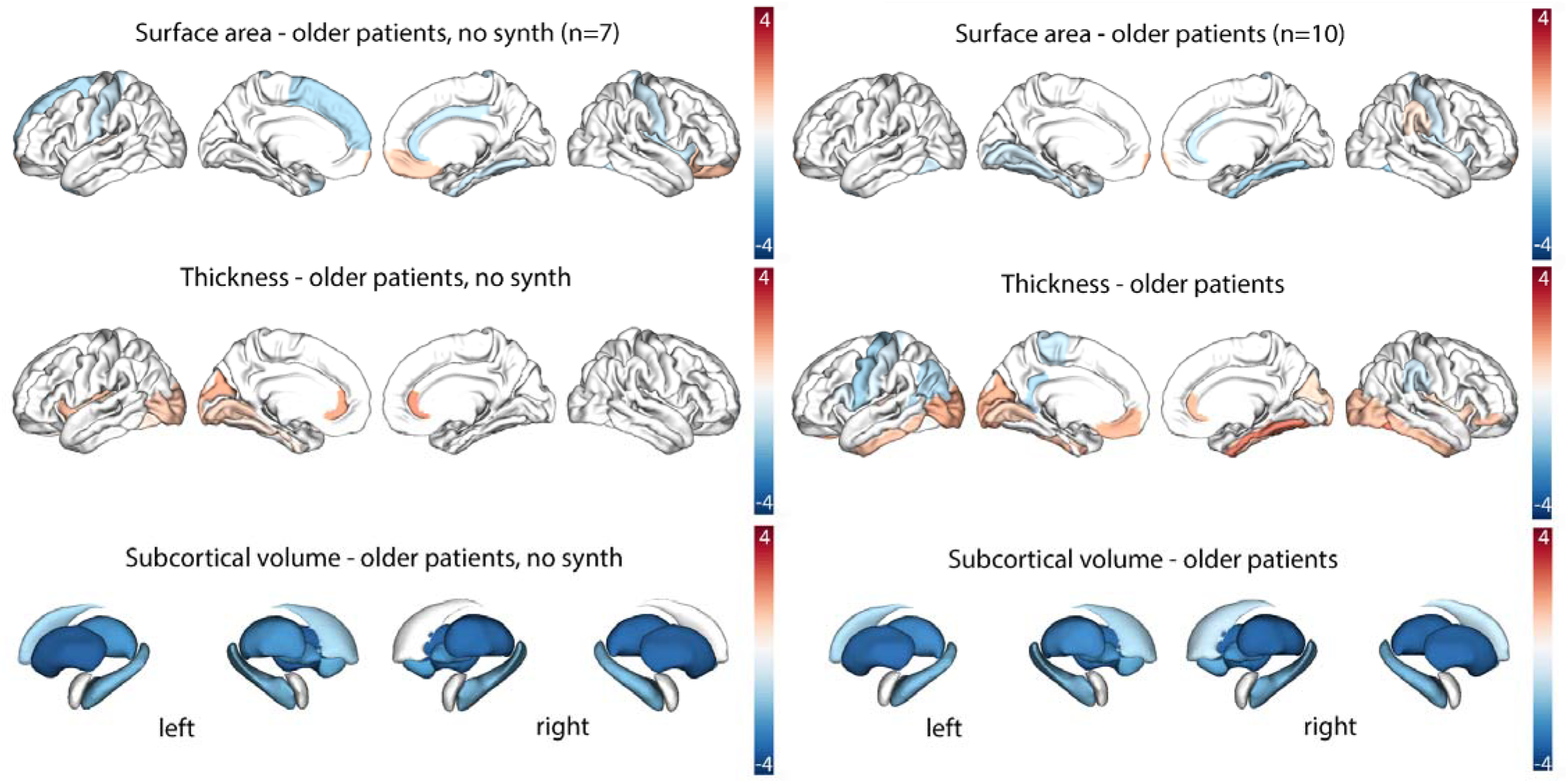
Average Z-scores for only native resolution (left) and the full sample (right) of older patients. Regions where the Z-score differed from zero (*p*<.05, uncorrected). Marginally significant regions (.0042<*p*<.05) are included to display trends. Differences in cortical surface area (top), cortical thickness (middle), and subcortical volume are shown for the older patients only, with native resolution only scans in the left panel and native resolution (n=6) plus enhanced clinical scans (n=4) in the right panel. Color corresponds to the average Z-score across the group for a given region, with negative Z-scores in blue and positive Z-scores in red. Left in image is left in brain.

**Supplementary Table 1.**
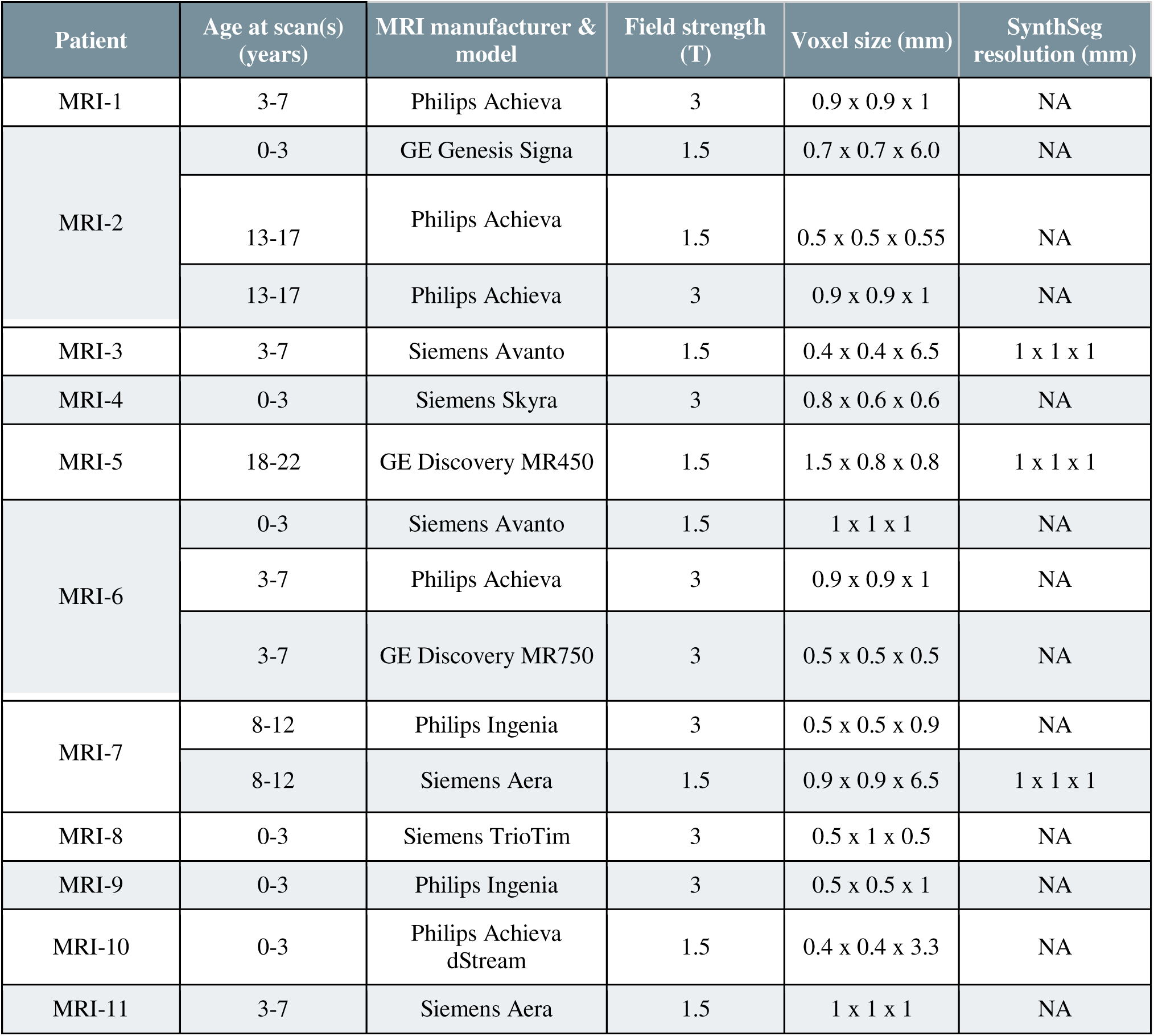
Scan parameters.

**Supplementary Table 2.**
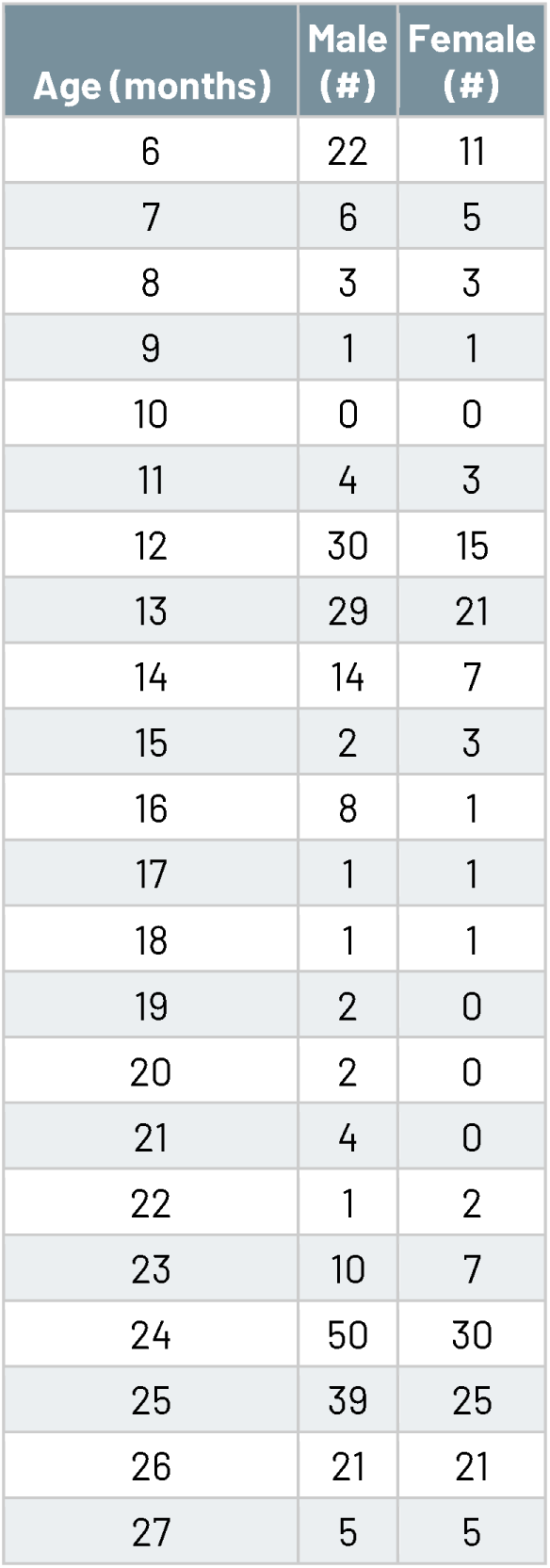
Data used for the infant normative database by participant age (in months) and sex. Data from 417 infants were pulled from the NIMH Data Archive (NDA). These include only participants designated as “typically developing” or “healthy control” by the contributing study and only those with scans that produced high quality FreeSurfer segmentations.

**Supplementary Table 3.**
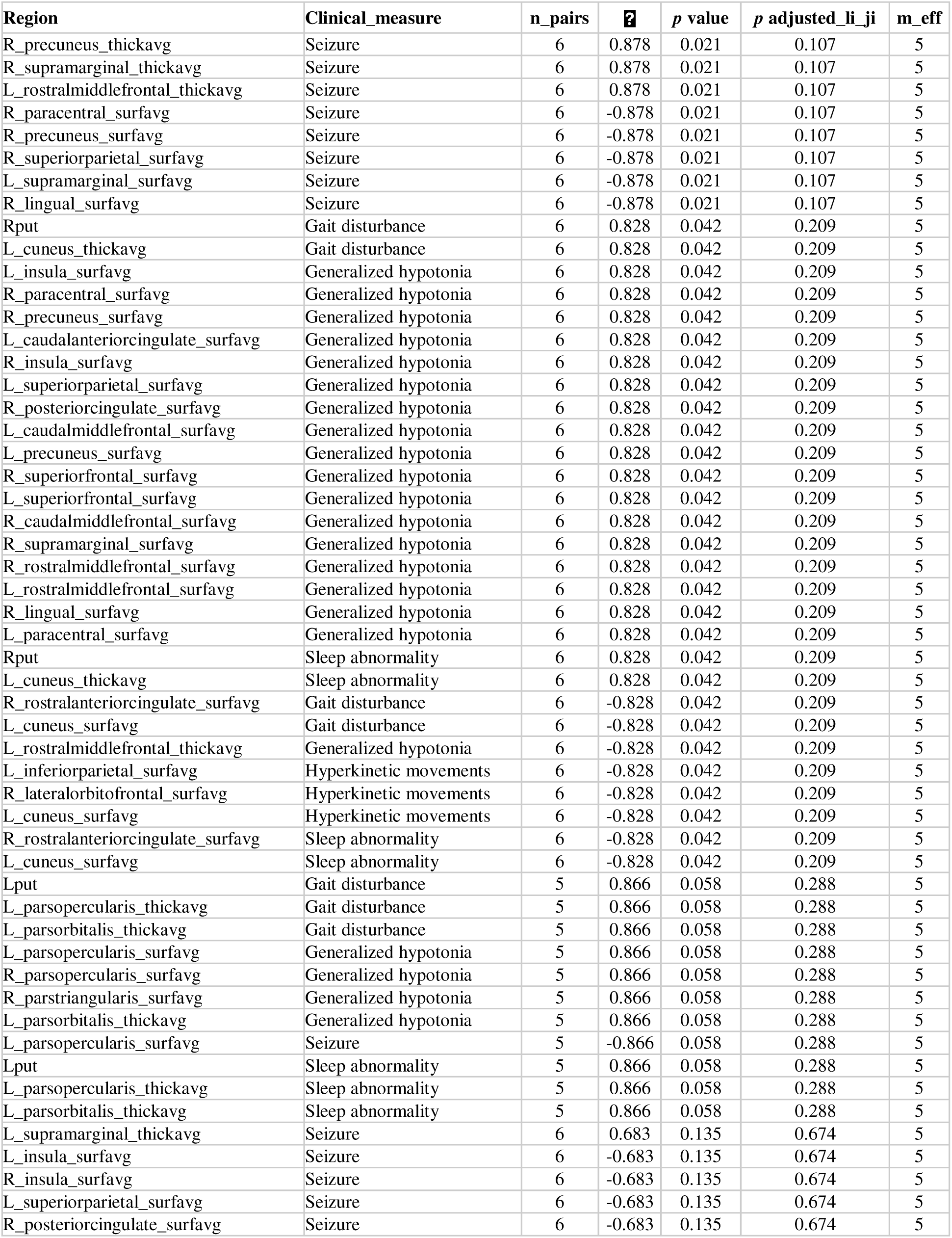
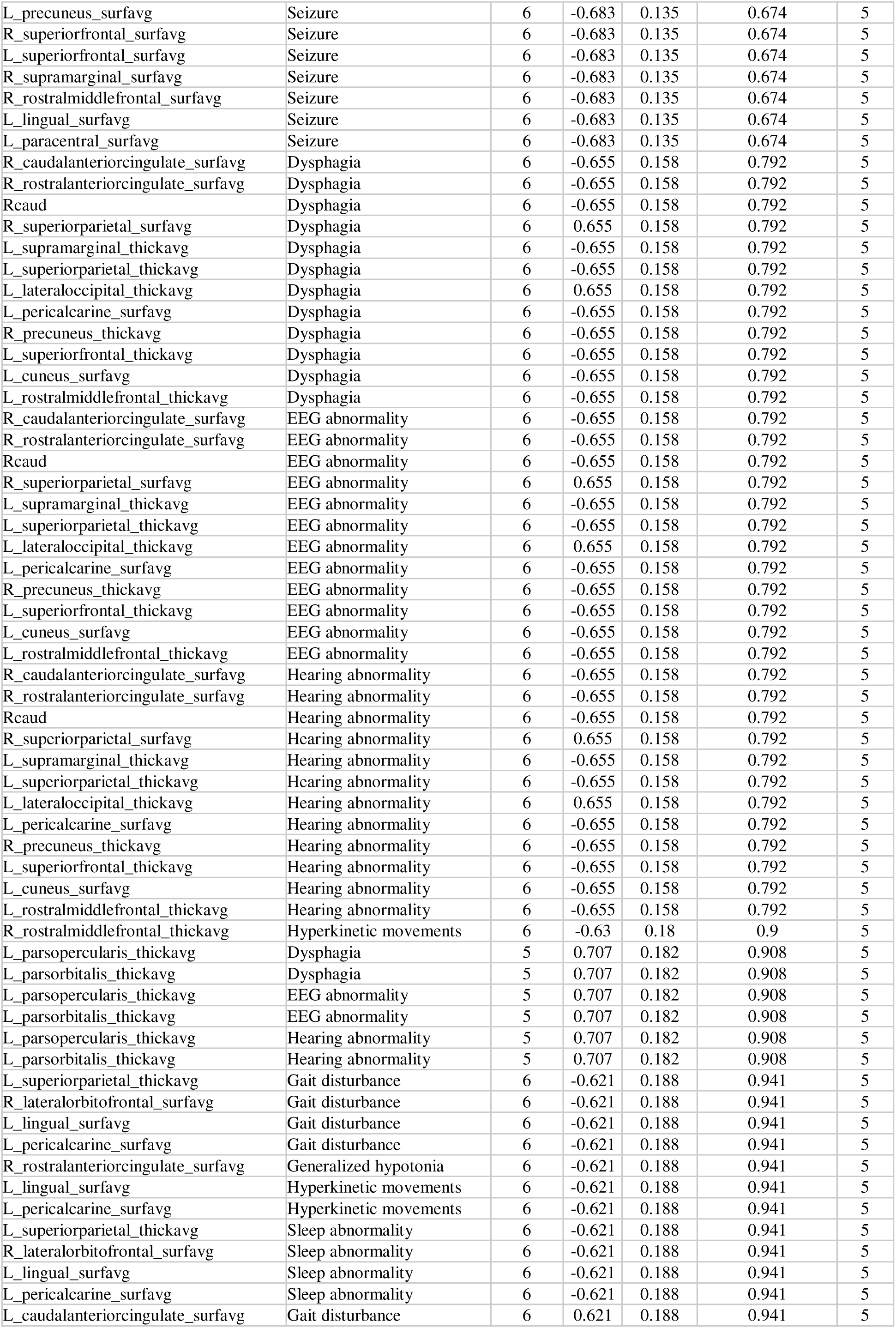
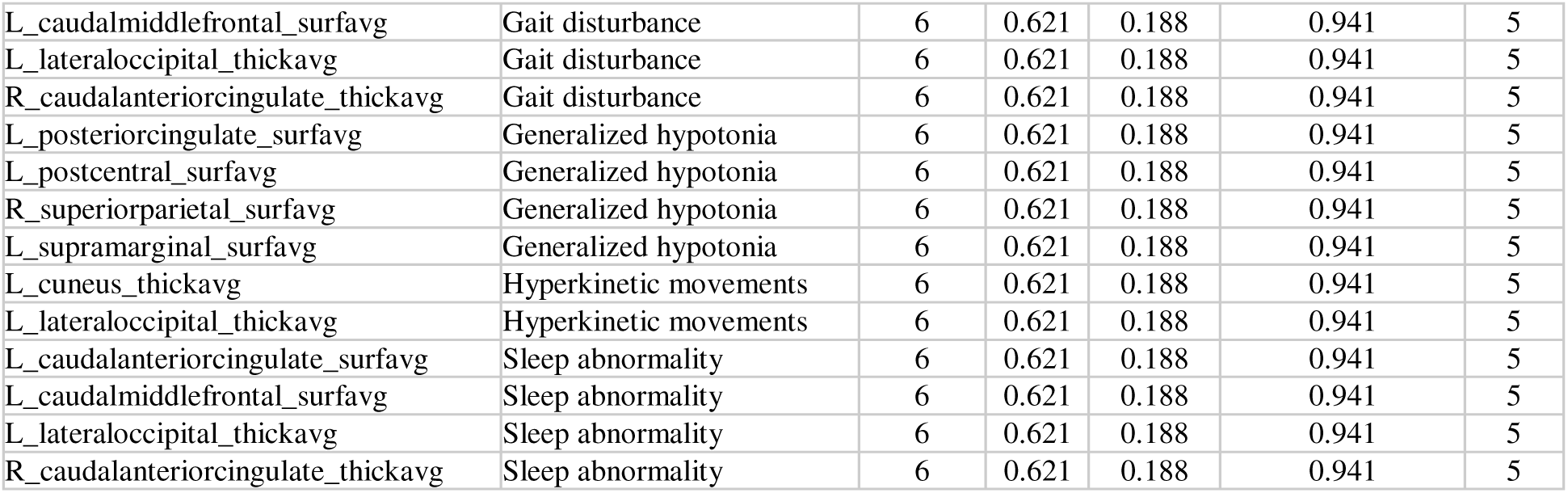
Complete clinical associations in the younger patient group. Among the regions where the mean Z-score differed from 0 with *p*<.10, we examined associations with 9 clinical measures (presence or absence). For each pair, the number of patients included, Spearman ⍰ (rho), raw *p*, adjusted *p* using the Li & Ji method, and the effective number of tests calculated by the Li & Ji method.

**Supplementary Table 4.**
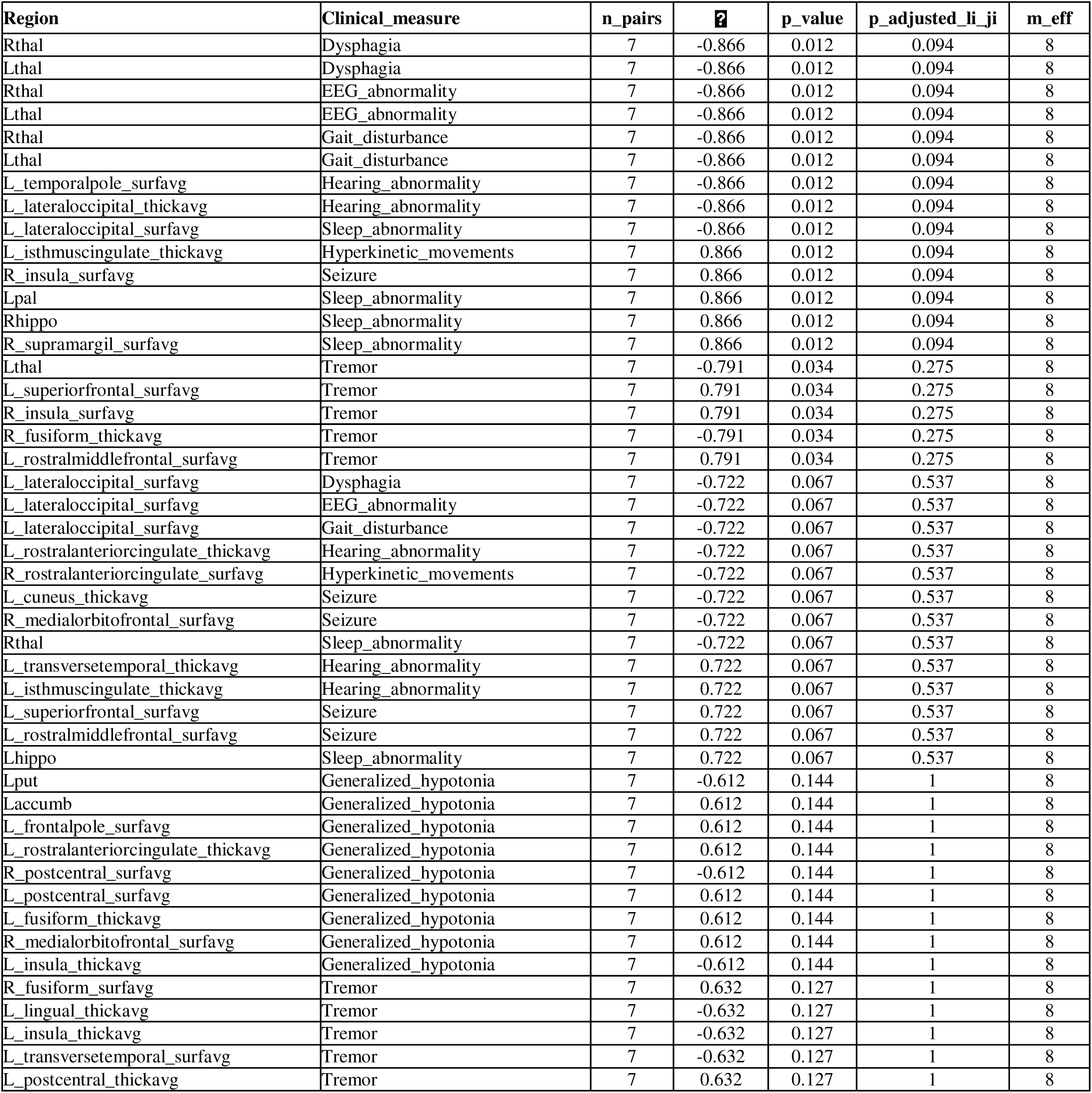
Complete clinical associations in the older patient group. Among the regions where the mean Z-score differed from 0 with *p*<.10, we examined associations with 9 clinical measures (presence or absence). For each pair, the number of patients included, Spearman ⍰ (rho), raw *p*, adjusted *p* using the Li & Ji method, and the effective number of tests calculated by the Li & Ji method.

---

**Table 1.**
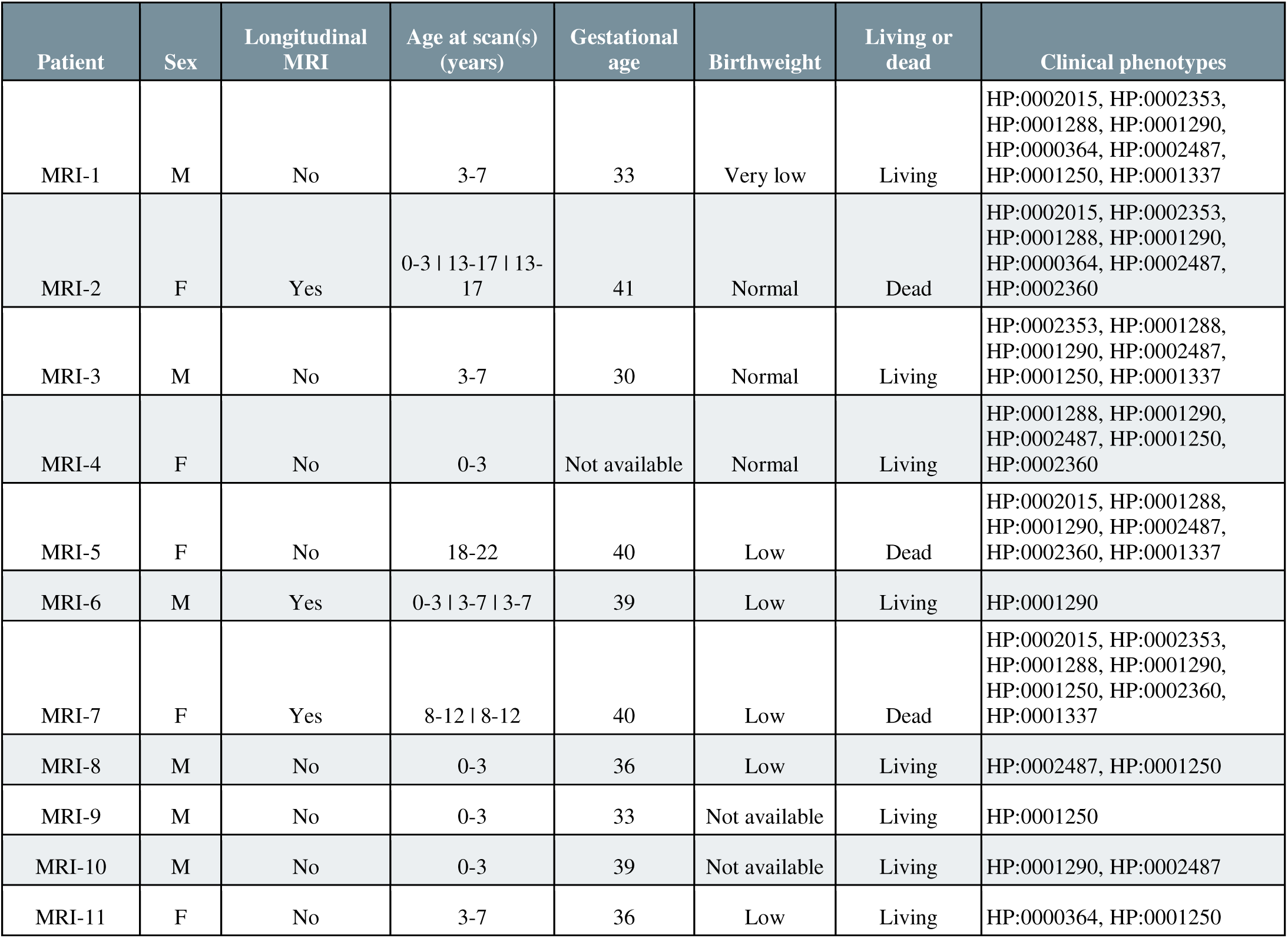
Patient characteristics. Demographic and clinical information for the 11 patients, including sex, whether longitudinal MRI was available, age at scan(s) in years, gestational age (in weeks), birthweight (very low [<1.5kg], low [1.5kg-2.5kg], and normal [>2.5kg]), whether patient was living as of 2023 or age at death (in years), and clinical phenotypes in HPO IDs: HP:0002015 - Dysphagia, HP:0002353 - EEG abnormality, HP:0001288 - Gait disturbance, HP:0001290 - Generalized hypotonia, HP:0000364 - Hearing abnormality, HP:0002487 - Hyperkinetic movements, HP:0001250 - Seizure, HP:0002360 - Sleep abnormality, HP:0001337 - Tremor.

| Patient | Sex | Longitudinal MRI | Age at scan(s) (years) | Gestational age | Birthweight | Living or dead | Clinical phenotypes |
| --- | --- | --- | --- | --- | --- | --- | --- |
| MRI-1 | M | No | 3-7 | 33 | Very low | Living | HP:0002015, HP:0002353, HP:0001288, HP:0001290, HP:0000364, HP:0002487, HP:0001250, HP:0001337 |
| MRI-2 | F | Yes | 0-3 \| 13-17 \| 13-17 | 41 | Normal | Dead | HP:0002015, HP:0002353, HP:0001288, HP:0001290, HP:0000364, HP:0002487, HP:0002360 |
| MRI-3 | M | No | 3-7 | 30 | Normal | Living | HP:0002353, HP:0001288, HP:0001290, HP:0002487, HP:0001250, HP:0001337 |
| MRI-4 | F | No | 0-3 | Not available | Normal | Living | HP:0001288, HP:0001290, HP:0002487, HP:0001250, HP:0002360 |
| MRI-5 | F | No | 18-22 | 40 | Low | Dead | HP:0002015, HP:0001288, HP:0001290, HP:0002487, HP:0002360, HP:0001337 |
| MRI-6 | M | Yes | 0-3 \| 3-7 \| 3-7 | 39 | Low | Living | HP:0001290 |
| MRI-7 | F | Yes | 8-12 \| 8-12 | 40 | Low | Dead | HP:0002015, HP:0002353, HP:0001288, HP:0001290, HP:0001250, HP:0002360, HP:0001337 |
| MRI-8 | M | No | 0-3 | 36 | Low | Living | HP:0002487, HP:0001250 |
| MRI-9 | M | No | 0-3 | 33 | Not available | Living | HP:0001250 |
| MRI-10 | M | No | 0-3 | 39 | Not available | Living | HP:0001290, HP:0002487 |
| MRI-11 | F | No | 3-7 | 36 | Low | Living | HP:0000364, HP:0001250 |

