## Supplement for "MRI Characterization of Structural Brain Abnormalities in NGLY1 Deficiency"

**Supplementary Table 4.** **Complete clinical associations in the older patient group.** Among the regions where the mean Z-score differed from 0 with *p*<.10, we examined associations with 9 clinical measures (presence or absence). For each pair, the number of patients included, Spearman ρ (rho), raw *p*, adjusted *p* using the Li & Ji method, and the effective number of tests calculated by the Li & Ji method.

**Supplementary Table 1.** Scan parameters.

| **Patient** | **Age at scan(s) (within range)** | **MRI manufacturer & model** | **Field strength (T)** | **Voxel size (mm)** | **SynthSeg resolution (mm)** |
| --- | --- | --- | --- | --- | --- |
| MRI-1 | 3-7 | Philips Achieva | 3 | 0.9 x 0.9 x 1 | NA |
| MRI-2 | 0-3 | GE Genesis Signa | 1.5 | 0.7 x 0.7 x 6.0 | NA |
|  | 13-17 | Philips Achieva | 1.5 | 0.5 x 0.5 x 0.55 | NA |
|  | 13-17 | Philips Achieva | 3 | 0.9 x 0.9 x 1 | NA |
| MRI-3 | 3-7 | Siemens Avanto | 1.5 | 0.4 x 0.4 x 6.5 | 1 x 1 x 1 |
| MRI-4 | 0-3 | Siemens Skyra | 3 | 0.8 x 0.6 x 0.6 | NA |
| MRI-5 | 18-22 | GE Discovery MR450 | 1.5 | 1.5 x 0.8 x 0.8 | 1 x 1 x 1 |
| MRI-6 | 0-3 | Siemens Avanto | 1.5 | 1 x 1 x 1 | NA |
|  | 3-7 | Philips Achieva | 3 | 0.9 x 0.9 x 1 | NA |
|  | 3-7 | GE Discovery MR750 | 3 | 0.5 x 0.5 x 0.5 | NA |
| MRI-7 | 8-12 | Philips Ingenia | 3 | 0.5 x 0.5 x 0.9 | NA |
|  | 8-12 | Siemens Aera | 1.5 | 0.9 x 0.9 x 6.5 | 1 x 1 x 1 |
| MRI-8 | 0-3 | Siemens TrioTim | 3 | 0.5 x 1 x 0.5 | NA |
| MRI-9 | 0-3 | Philips Ingenia | 3 | 0.5 x 0.5 x 1 | NA |
| MRI-10 | 0-3 | Philips Achieva dStream | 1.5 | 0.4 x 0.4 x 3.3 | NA |
| MRI-11 | 3-7 | Siemens Aera | 1.5 | 1 x 1 x 1 | NA |

**Supplementary Table 2.** Data used for the infant normative database by participant age (in months) and sex. Data from 417 infants were pulled from the NIMH Data Archive (NDA). These include only participants designated as “typically developing” or “healthy control” by the contributing study and only those with scans that produced high quality FreeSurfer segmentations.

| **Age (months)** | **Male (#)** | **Female (#)** |
| --- | --- | --- |
| 6 | 22 | 11 |
| 7 | 6 | 5 |
| 8 | 3 | 3 |
| 9 | 1 | 1 |
| 10 | 0 | 0 |
| 11 | 4 | 3 |
| 12 | 30 | 15 |
| 13 | 29 | 21 |
| 14 | 14 | 7 |
| 15 | 2 | 3 |
| 16 | 8 | 1 |
| 17 | 1 | 1 |
| 18 | 1 | 1 |
| 19 | 2 | 0 |
| 20 | 2 | 0 |
| 21 | 4 | 0 |
| 22 | 1 | 2 |
| 23 | 10 | 7 |
| 24 | 50 | 30 |
| 25 | 39 | 25 |
| 26 | 21 | 21 |
| 27 | 5 | 5 |

**Supplementary Figure 1. Normative trajectories of subcortical volume for ages 6-27 months.** Normative trajectories created from 417 infants in the NDA, separated by sex. Mean, 1 standard deviation (SD), and 2SD curves are shown for males (blue) and females (red).

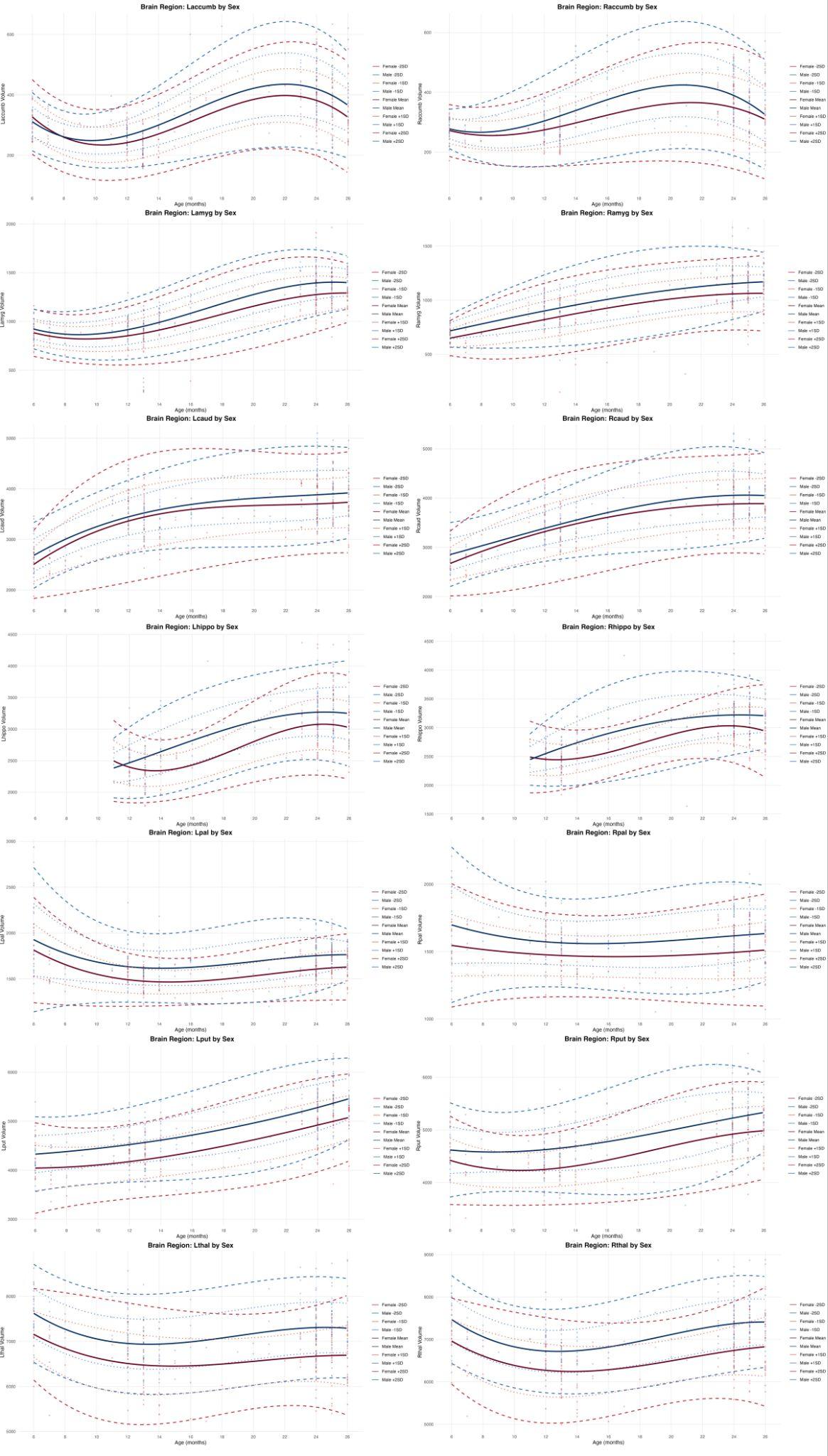

**Supplementary Figure 2. Normative trajectories of cortical thickness (cingulate, insula, and occipital lobes) for ages 6-27 months.** Normative trajectories created from 417 infants, separated by sex. Mean, 1 standard deviation (SD), and 2SD curves are shown for males (blue) and females (red).

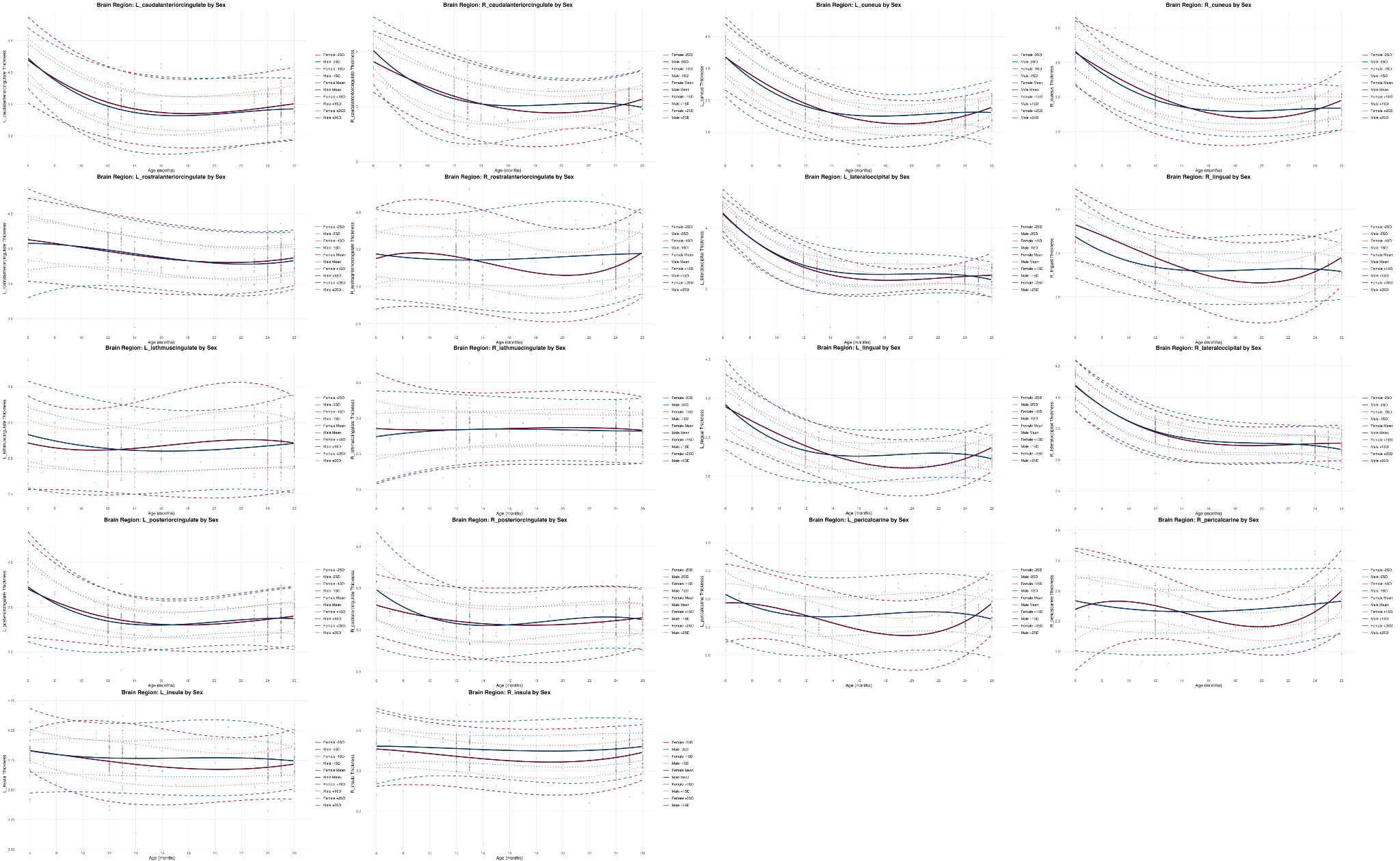

**Supplementary Figure 3. Normative trajectories of cortical thickness (frontal lobe) for ages 6-27 months.** Normative trajectories created from 417 infants, separated by sex. Mean, 1 standard deviation (SD), and 2SD curves are shown for males (blue) and females (red).

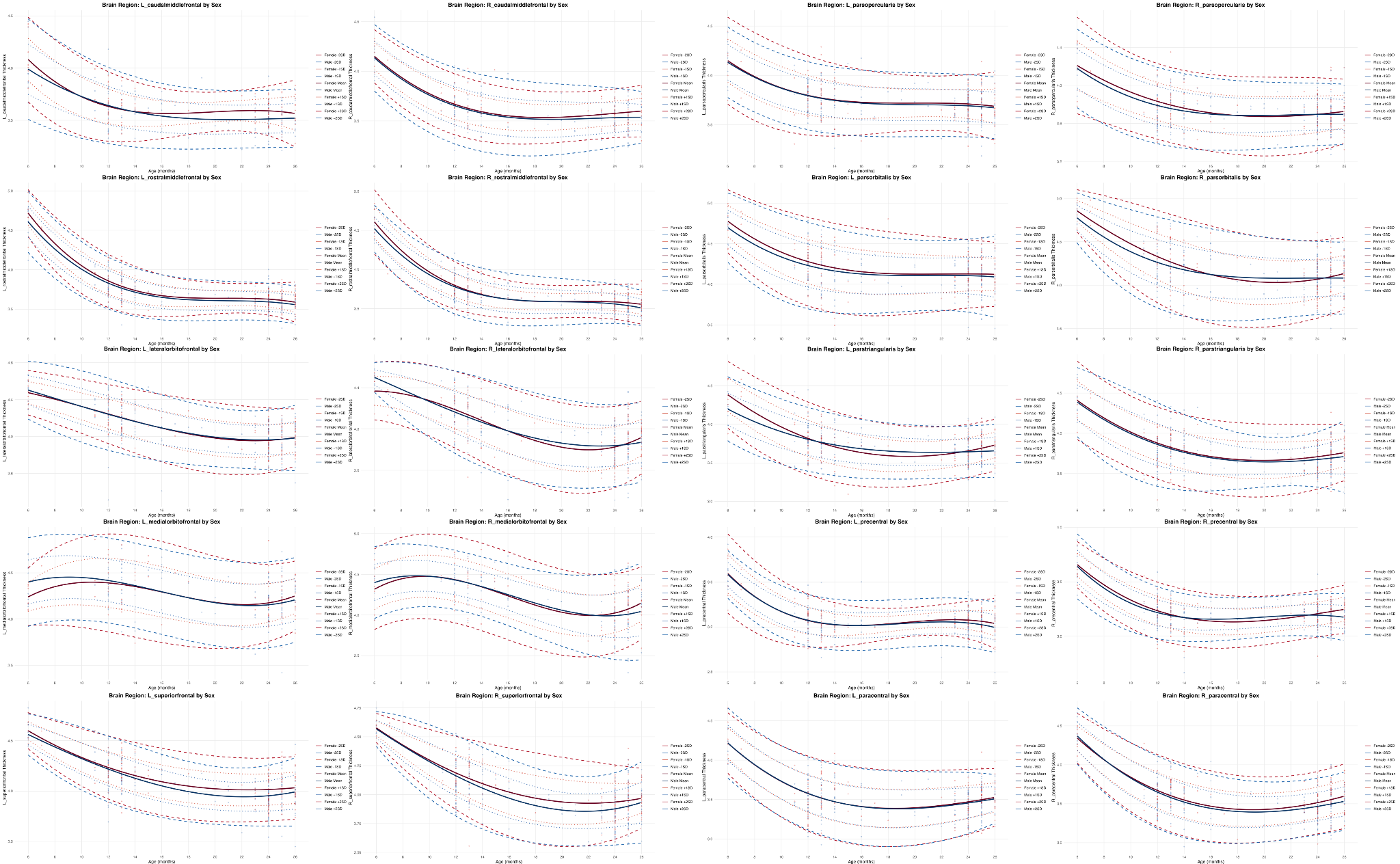

**Supplementary Figure 4. Normative trajectories of cortical thickness (parietal and temporal lobes) for ages 6-27 months.** Normative trajectories created from 417 infants, separated by sex. Mean, 1 standard deviation (SD), and 2SD curves are shown for males (blue) and females (red).

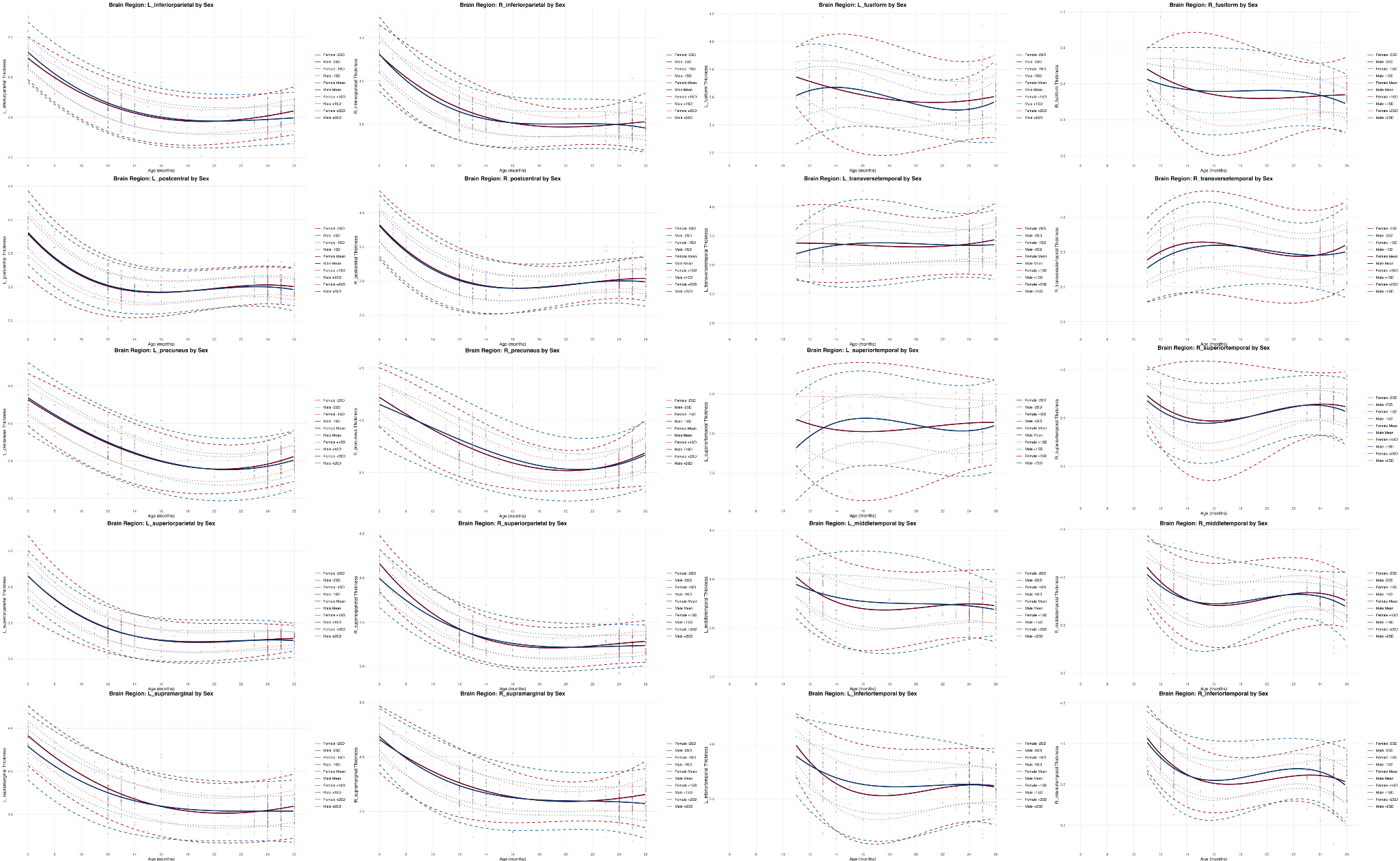

**Supplementary Figure 5. Normative trajectories of cortical surface area (cingulate, insula, and occipital lobes) for ages 6-27 months.** Normative trajectories created from 417 infants, separated by sex. Mean, 1 standard deviation (SD), and 2SD curves are shown for males (blue) and females (red).

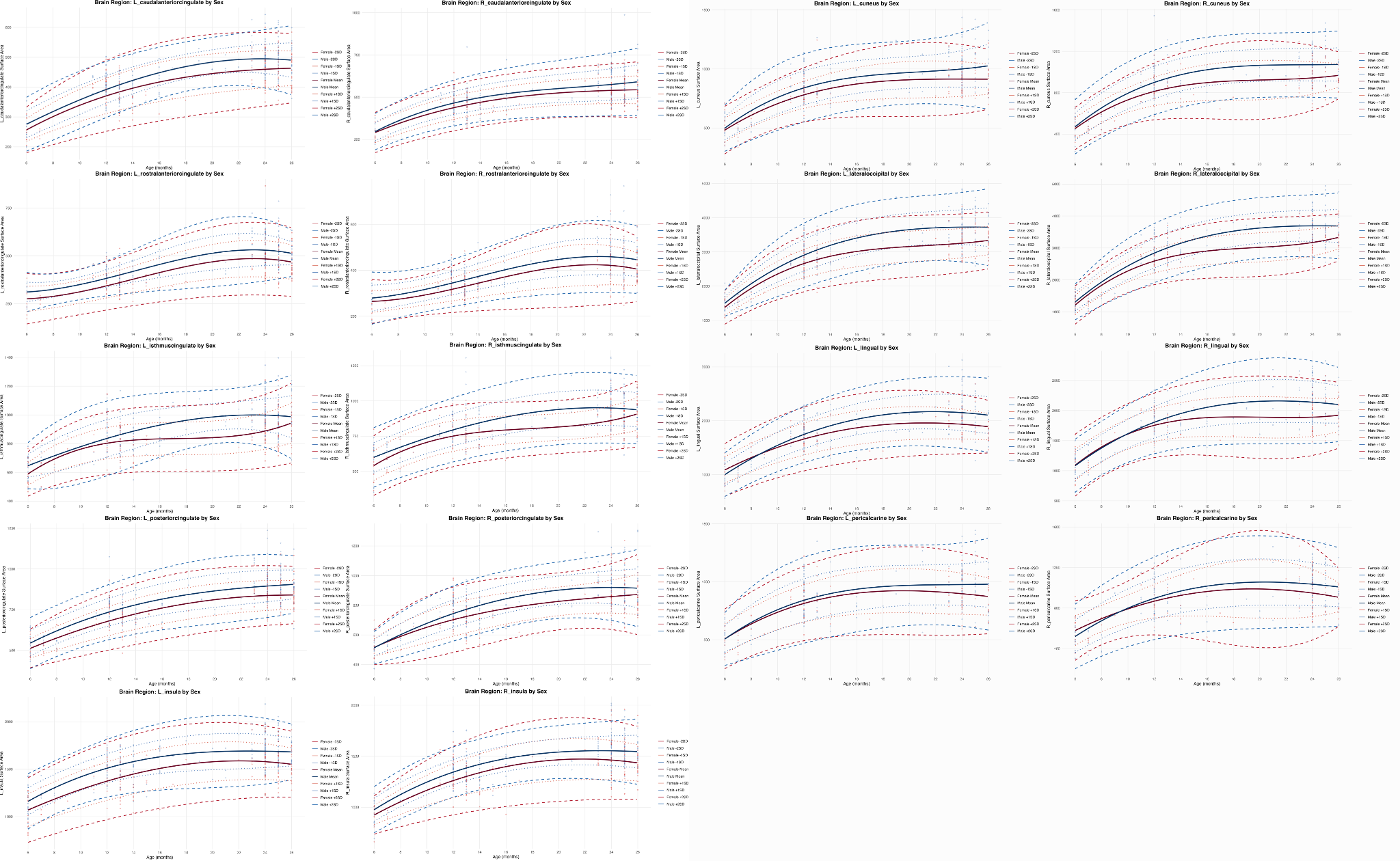

**Supplementary Figure 6. Normative trajectories of cortical surface area (frontal lobe) for ages 6-27 months.** Normative trajectories created from 417 infants, separated by sex. Mean, 1 standard deviation (SD), and 2SD curves are shown for males (blue) and females (red).

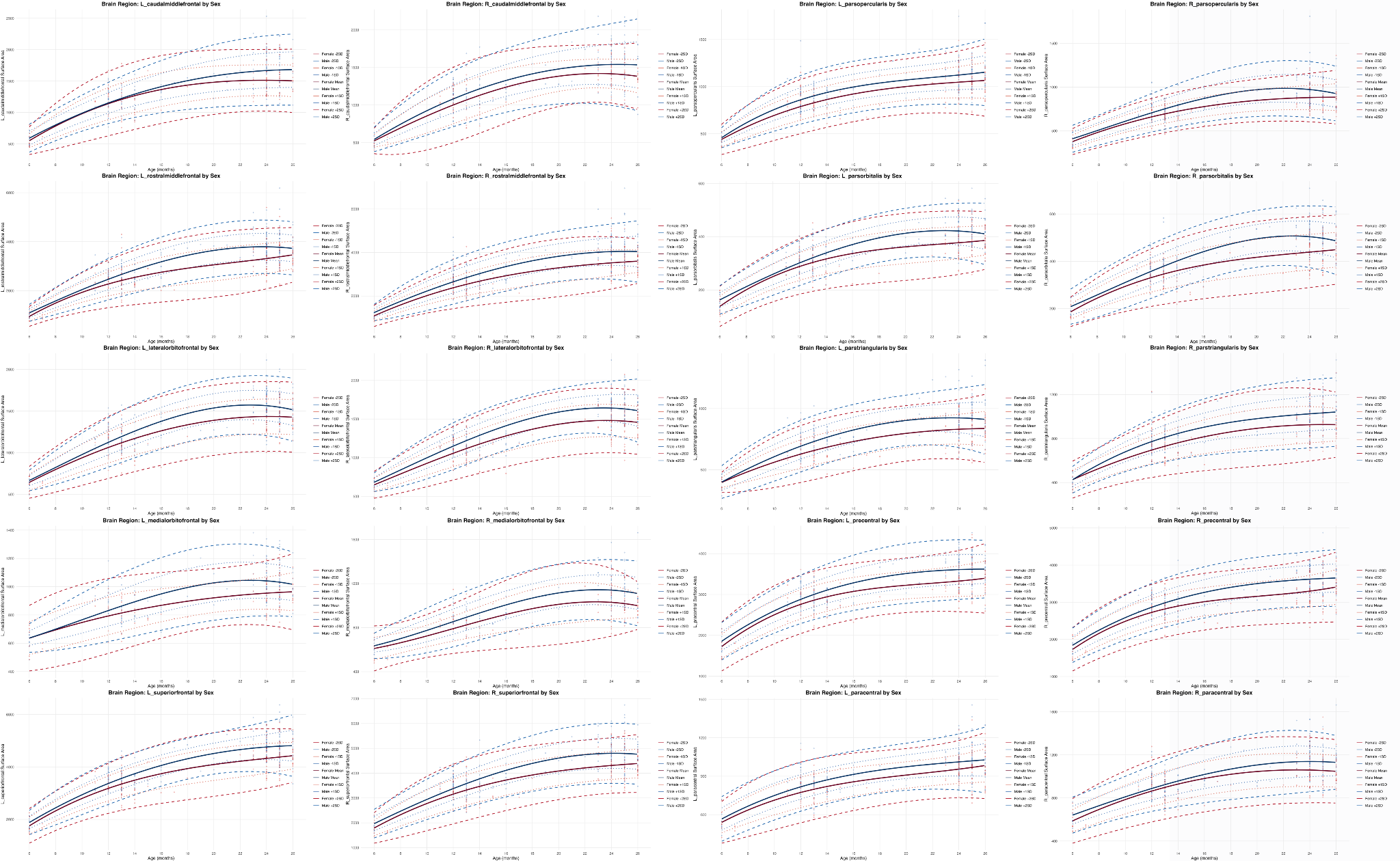

**Supplementary Figure 7. Normative trajectories of cortical surface area (parietal and temporal lobes) for ages 6-27 months.** Normative trajectories created from 417 infants, separated by sex. Mean, 1 standard deviation (SD), and 2SD curves are shown for males (blue) and females (red).

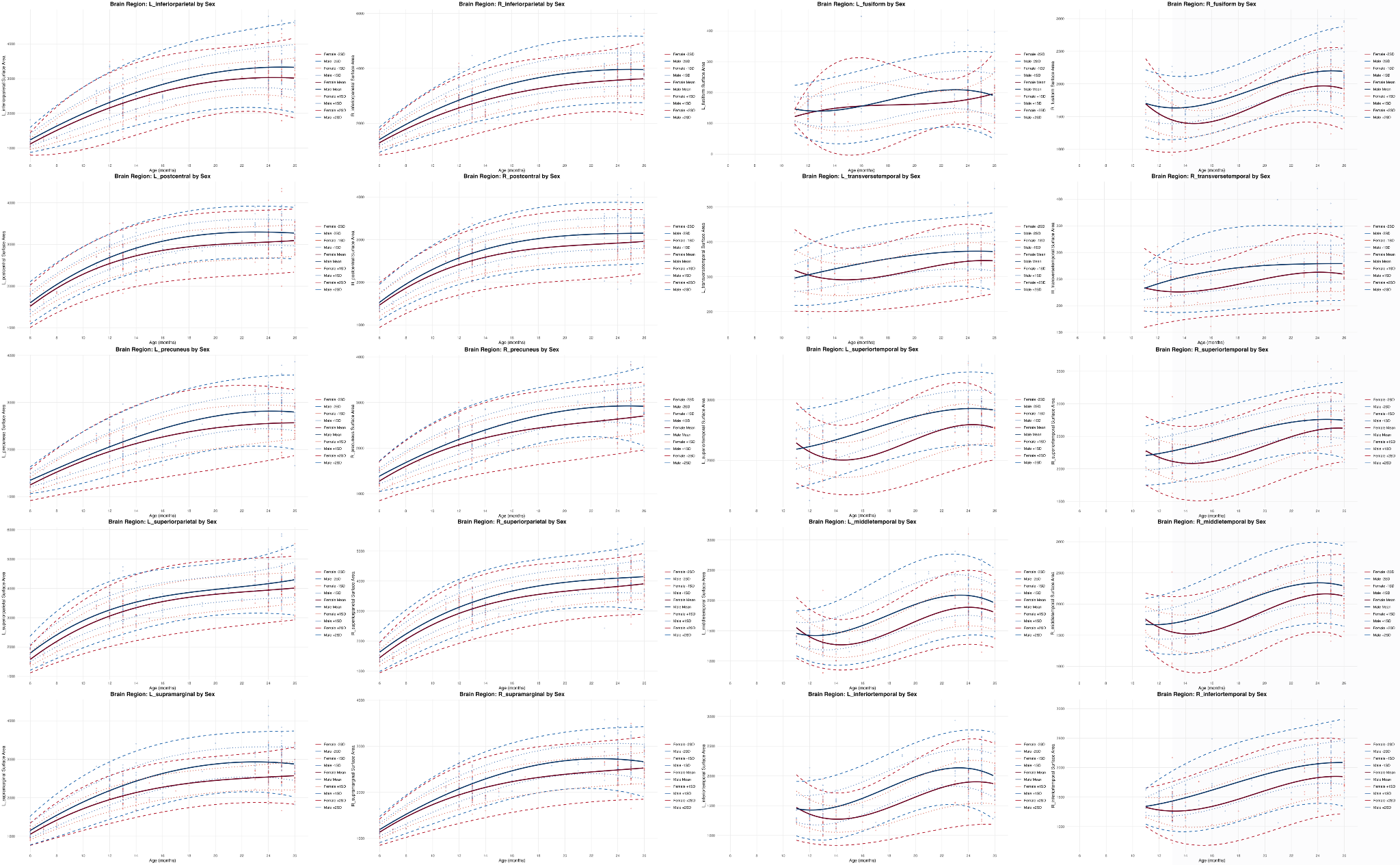

**Supplementary Figure 8.** Visualization of the cortical morphometry metrics examined in this paper. The pial boundary for an individual gyrus is outlined in the coronal pop out in white, with the boundary between gray and white matter outlined in black. The distance between these, shown in the red lines, is the cortical thickness. The surface area is the area of the gray matter surface, as shown in blue. Volume is the gray matter volume of a region, as shown in green. Curvature is local folding of the cortex and is defined by the radius of the sphere that fits the curve. Mean curvature is the average of the first two principal components of curvature for a region, which are orthogonal to each other.

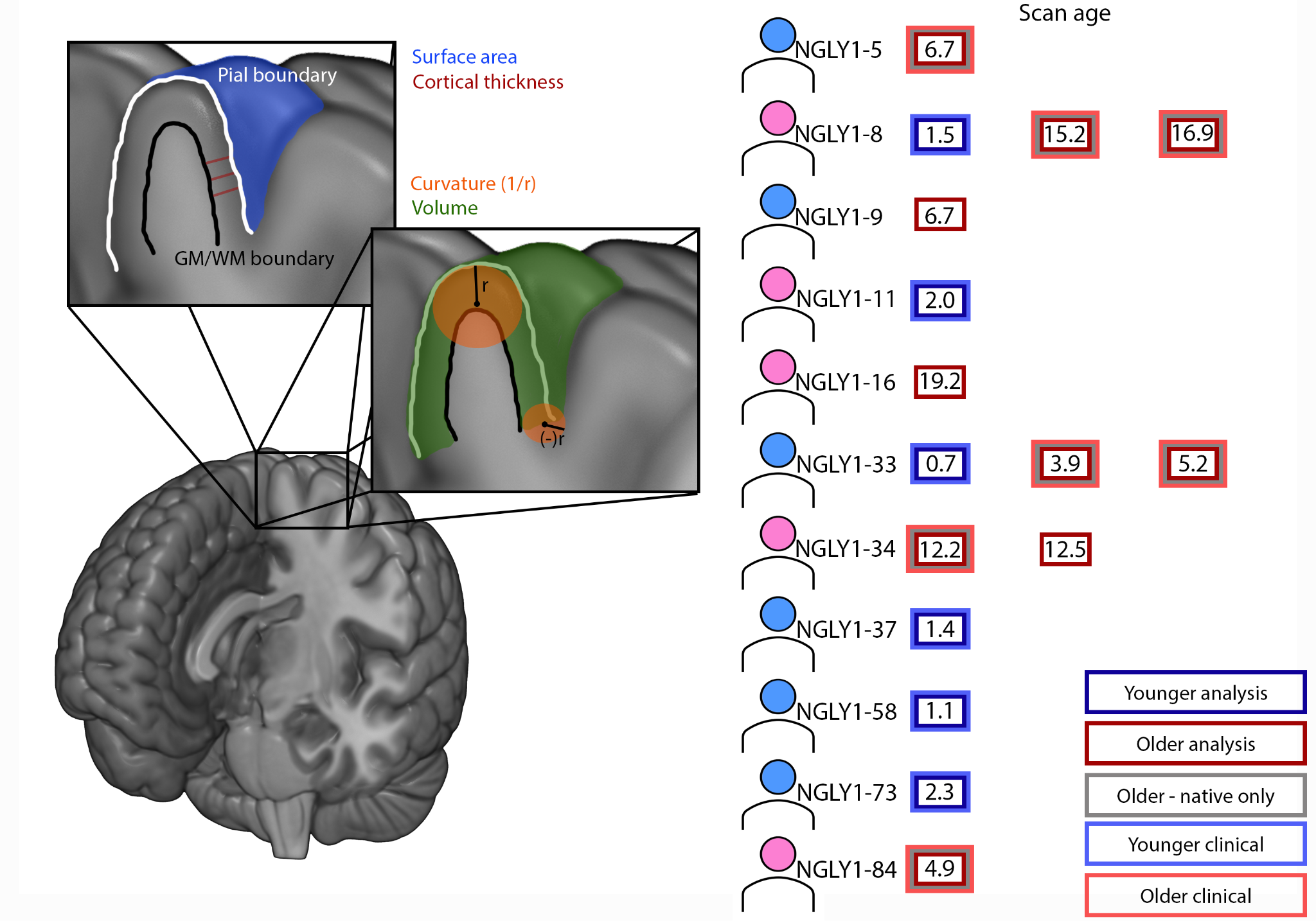

**Supplementary Figure 9. Subcortical volume Z-scores in older patients (real only and combined).** Boxplots display Z-scores for each subcortical structure (top: only native resolution scans, bottom: native and SynthSR enhanced scans) relative to age- and sex-matched normative data. Box elements represent median, interquartile range (IQR), and whiskers extend to minimum and maximum excluding outliers; points beyond whiskers are outliers. Symbols indicate significant deviations from normative means: ^#^p<.05 after multiple comparisons correction (Li & Ji); *p<.05 uncorrected.

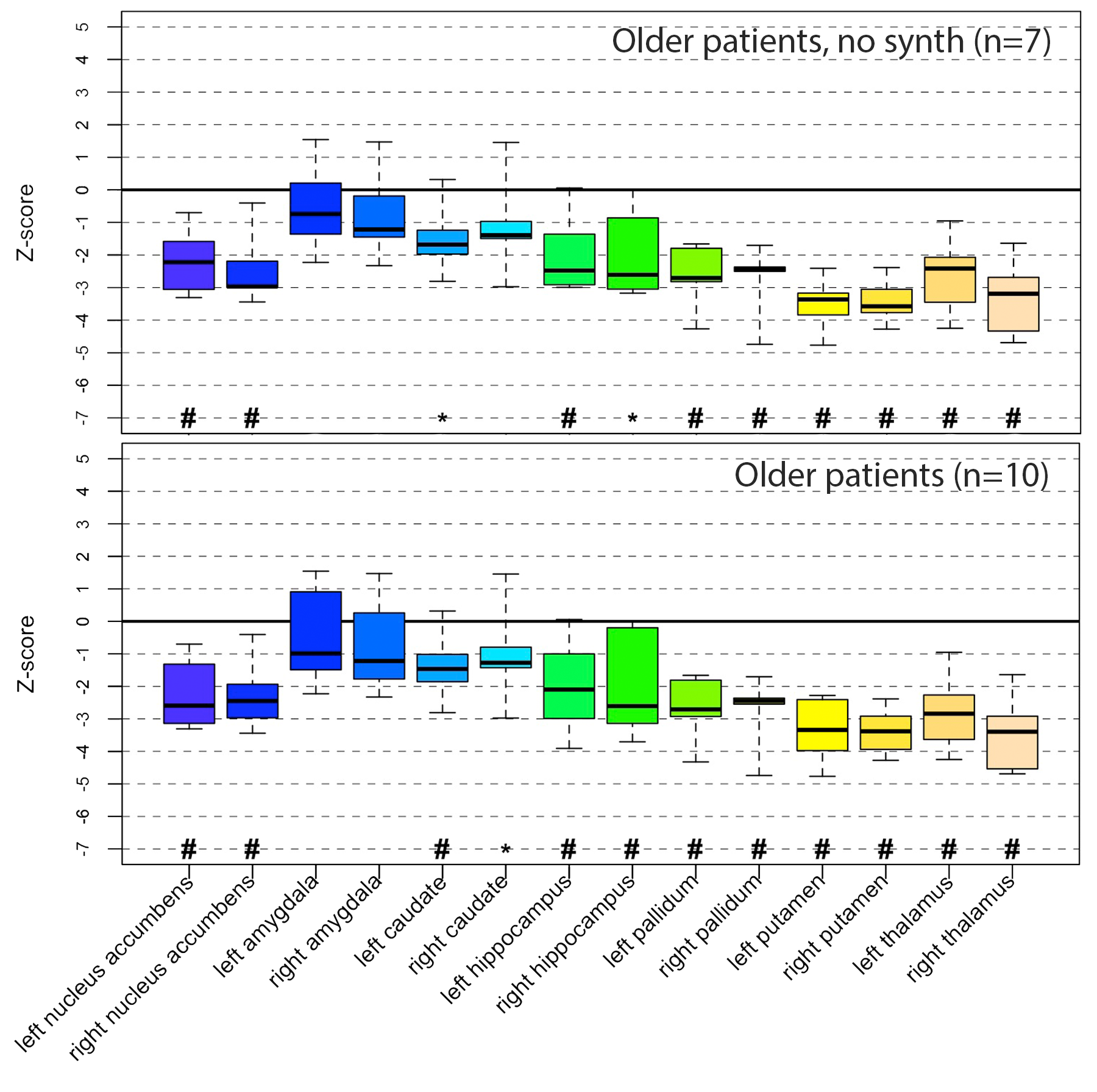

**Supplementary Figure 10. Cortical thickness Z-scores in older patients (native resolution only [upper] and combined native and SynthSR [lower]).** Boxplots display Z-scores for each cortical region (top: only native resolution scans, bottom: native and SynthSR enhanced scans) relative to age- and sex-matched normative data. Box elements represent median, interquartile range (IQR), and whiskers extend to minimum and maximum excluding outliers; points beyond whiskers are outliers. Symbols indicate significant deviations from normative means: ^#^p<.05 after multiple comparisons correction (Li & Ji); *p<.05 uncorrected.

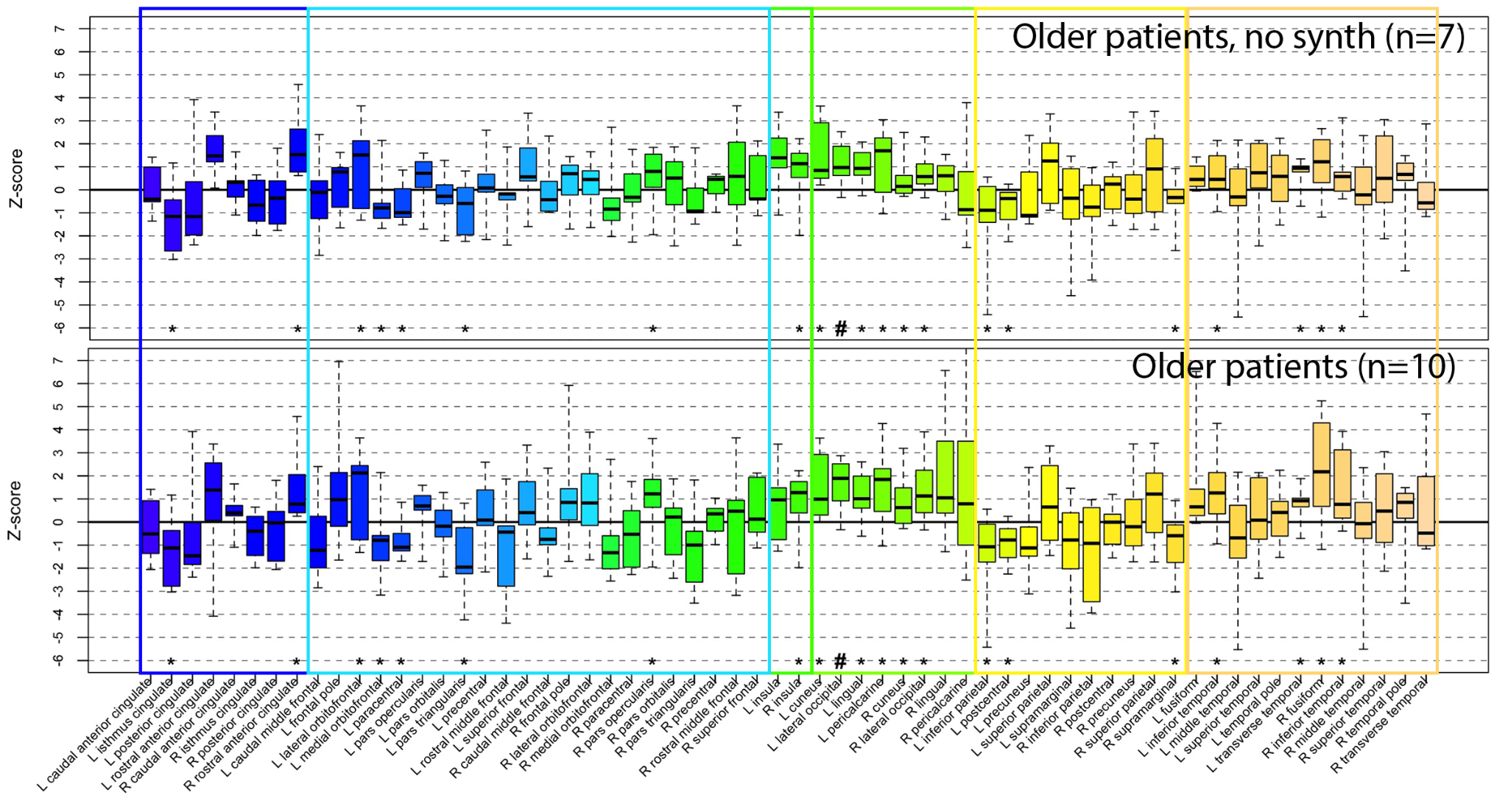

**Supplementary Figure 11. Cortical surface area Z-scores in older patients (native resolution only [upper] and combined native and SynthSR [lower]).** Boxplots display Z-scores for each cortical region (top: only native resolution scans, bottom: native and SynthSR enhanced scans) relative to age- and sex-matched normative data. Box elements represent median, interquartile range (IQR), and whiskers extend to minimum and maximum excluding outliers; points beyond whiskers are outliers. Symbols indicate significant deviations from normative means: ^#^p<.05 after multiple comparisons correction (Li & Ji); *p<.05 uncorrected.

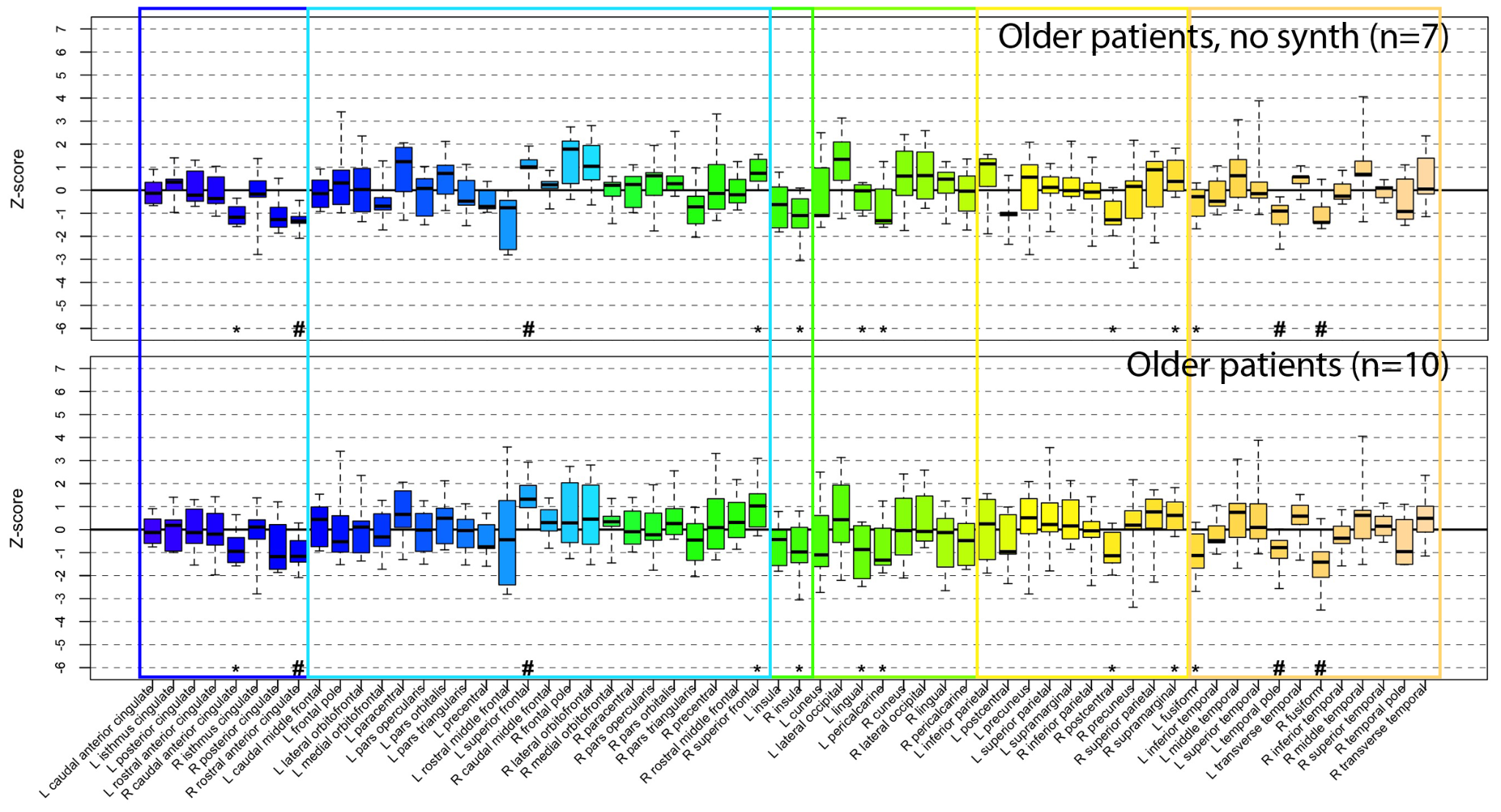

**Supplementary Figure 12. Average Z-scores for only native resolution (left) and the full sample (right) of older patients.** Regions where the Z-score differed from zero (*p*<.05, uncorrected). Marginally significant regions (.0042<*p*<.05) are included to display trends. Differences in cortical surface area (top), cortical thickness (middle), and subcortical volume are shown for the older patients only, with native resolution only scans in the left panel and native resolution (n=6) plus enhanced clinical scans (n=4) in the right panel. Color corresponds to the average Z-score across the group for a given region, with negative Z-scores in blue and positive Z-scores in red. Left in image is left in brain.

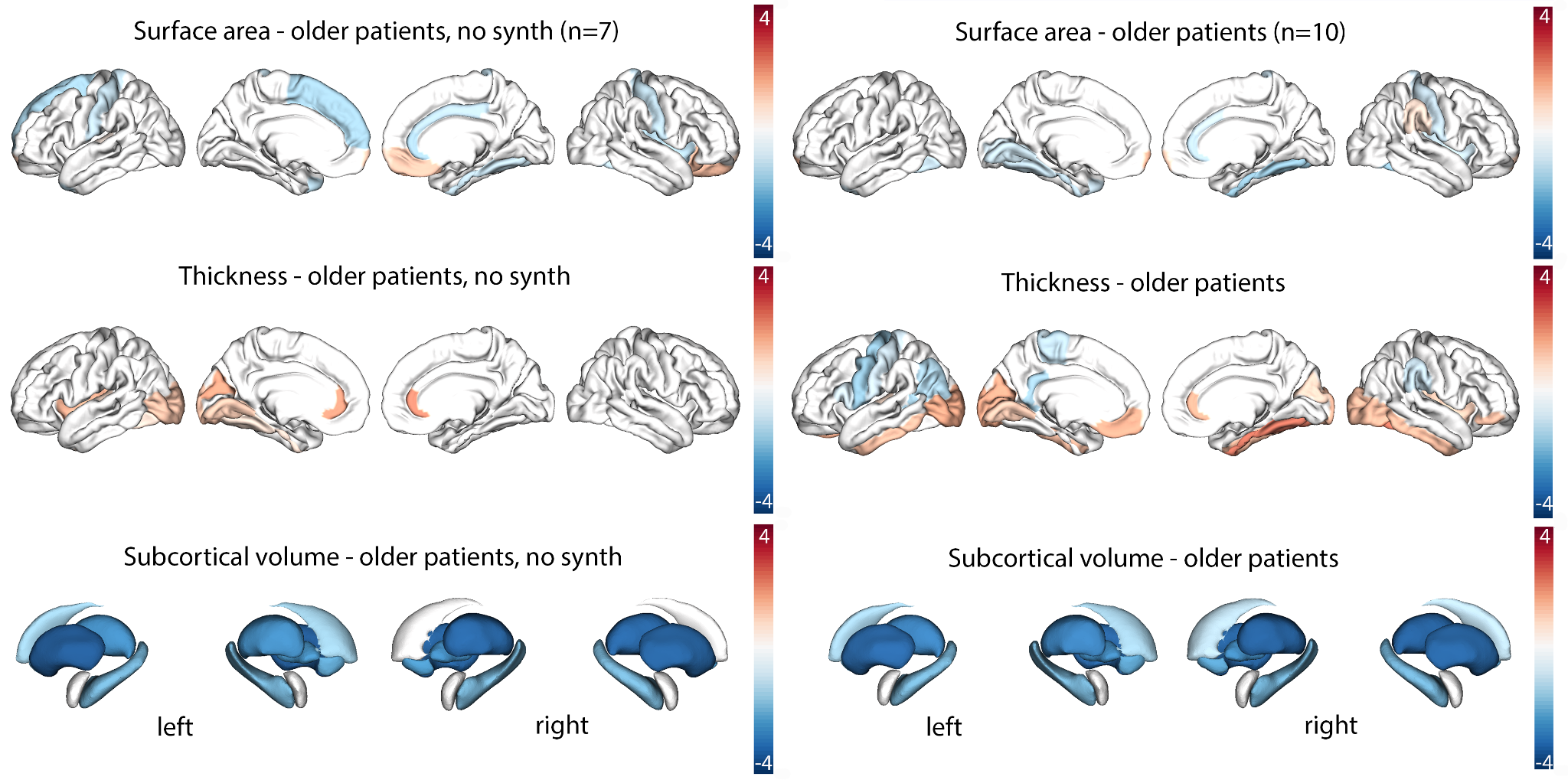

**Supplementary Table 3.** **Complete clinical associations in the younger patient group.** Among the regions where the mean Z-score differed from 0 with *p*<.10, we examined associations (presence or absence) with 9 clinical measures . For each region-clinical phenotype pair, the number of patients included, Spearman ρ (rho), raw *p-*value, adjusted *p-*value using the Li & Ji method, and the effective number of tests calculated by the Li & Ji method are reported.

| **Region** | **Clinical_measure** | **n_pairs** | **ρ** | ***p* value** | ***p* adjusted_li_ji** | **m_eff** |
| --- | --- | --- | --- | --- | --- | --- |
| R_precuneus_thickavg | Seizure | 6 | 0.878 | 0.021 | 0.107 | 5 |
| R_supramarginal_thickavg | Seizure | 6 | 0.878 | 0.021 | 0.107 | 5 |
| L_rostralmiddlefrontal_thickavg | Seizure | 6 | 0.878 | 0.021 | 0.107 | 5 |
| R_paracentral_surfavg | Seizure | 6 | -0.878 | 0.021 | 0.107 | 5 |
| R_precuneus_surfavg | Seizure | 6 | -0.878 | 0.021 | 0.107 | 5 |
| R_superiorparietal_surfavg | Seizure | 6 | -0.878 | 0.021 | 0.107 | 5 |
| L_supramarginal_surfavg | Seizure | 6 | -0.878 | 0.021 | 0.107 | 5 |
| R_lingual_surfavg | Seizure | 6 | -0.878 | 0.021 | 0.107 | 5 |
| Rput | Gait disturbance | 6 | 0.828 | 0.042 | 0.209 | 5 |
| L_cuneus_thickavg | Gait disturbance | 6 | 0.828 | 0.042 | 0.209 | 5 |
| L_insula_surfavg | Generalized hypotonia | 6 | 0.828 | 0.042 | 0.209 | 5 |
| R_paracentral_surfavg | Generalized hypotonia | 6 | 0.828 | 0.042 | 0.209 | 5 |
| R_precuneus_surfavg | Generalized hypotonia | 6 | 0.828 | 0.042 | 0.209 | 5 |
| L_caudalanteriorcingulate_surfavg | Generalized hypotonia | 6 | 0.828 | 0.042 | 0.209 | 5 |
| R_insula_surfavg | Generalized hypotonia | 6 | 0.828 | 0.042 | 0.209 | 5 |
| L_superiorparietal_surfavg | Generalized hypotonia | 6 | 0.828 | 0.042 | 0.209 | 5 |
| R_posteriorcingulate_surfavg | Generalized hypotonia | 6 | 0.828 | 0.042 | 0.209 | 5 |
| L_caudalmiddlefrontal_surfavg | Generalized hypotonia | 6 | 0.828 | 0.042 | 0.209 | 5 |
| L_precuneus_surfavg | Generalized hypotonia | 6 | 0.828 | 0.042 | 0.209 | 5 |
| R_superiorfrontal_surfavg | Generalized hypotonia | 6 | 0.828 | 0.042 | 0.209 | 5 |
| L_superiorfrontal_surfavg | Generalized hypotonia | 6 | 0.828 | 0.042 | 0.209 | 5 |
| R_caudalmiddlefrontal_surfavg | Generalized hypotonia | 6 | 0.828 | 0.042 | 0.209 | 5 |
| R_supramarginal_surfavg | Generalized hypotonia | 6 | 0.828 | 0.042 | 0.209 | 5 |
| R_rostralmiddlefrontal_surfavg | Generalized hypotonia | 6 | 0.828 | 0.042 | 0.209 | 5 |
| L_rostralmiddlefrontal_surfavg | Generalized hypotonia | 6 | 0.828 | 0.042 | 0.209 | 5 |
| R_lingual_surfavg | Generalized hypotonia | 6 | 0.828 | 0.042 | 0.209 | 5 |
| L_paracentral_surfavg | Generalized hypotonia | 6 | 0.828 | 0.042 | 0.209 | 5 |
| Rput | Sleep abnormality | 6 | 0.828 | 0.042 | 0.209 | 5 |
| L_cuneus_thickavg | Sleep abnormality | 6 | 0.828 | 0.042 | 0.209 | 5 |
| R_rostralanteriorcingulate_surfavg | Gait disturbance | 6 | -0.828 | 0.042 | 0.209 | 5 |
| L_cuneus_surfavg | Gait disturbance | 6 | -0.828 | 0.042 | 0.209 | 5 |
| L_rostralmiddlefrontal_thickavg | Generalized hypotonia | 6 | -0.828 | 0.042 | 0.209 | 5 |
| L_inferiorparietal_surfavg | Hyperkinetic movements | 6 | -0.828 | 0.042 | 0.209 | 5 |
| R_lateralorbitofrontal_surfavg | Hyperkinetic movements | 6 | -0.828 | 0.042 | 0.209 | 5 |
| L_cuneus_surfavg | Hyperkinetic movements | 6 | -0.828 | 0.042 | 0.209 | 5 |
| R_rostralanteriorcingulate_surfavg | Sleep abnormality | 6 | -0.828 | 0.042 | 0.209 | 5 |
| L_cuneus_surfavg | Sleep abnormality | 6 | -0.828 | 0.042 | 0.209 | 5 |
| Lput | Gait disturbance | 5 | 0.866 | 0.058 | 0.288 | 5 |
| L_parsopercularis_thickavg | Gait disturbance | 5 | 0.866 | 0.058 | 0.288 | 5 |
| L_parsorbitalis_thickavg | Gait disturbance | 5 | 0.866 | 0.058 | 0.288 | 5 |
| L_parsopercularis_surfavg | Generalized hypotonia | 5 | 0.866 | 0.058 | 0.288 | 5 |
| R_parsopercularis_surfavg | Generalized hypotonia | 5 | 0.866 | 0.058 | 0.288 | 5 |
| R_parstriangularis_surfavg | Generalized hypotonia | 5 | 0.866 | 0.058 | 0.288 | 5 |
| L_parsorbitalis_thickavg | Generalized hypotonia | 5 | 0.866 | 0.058 | 0.288 | 5 |
| L_parsopercularis_surfavg | Seizure | 5 | -0.866 | 0.058 | 0.288 | 5 |
| Lput | Sleep abnormality | 5 | 0.866 | 0.058 | 0.288 | 5 |
| L_parsopercularis_thickavg | Sleep abnormality | 5 | 0.866 | 0.058 | 0.288 | 5 |
| L_parsorbitalis_thickavg | Sleep abnormality | 5 | 0.866 | 0.058 | 0.288 | 5 |
| L_supramarginal_thickavg | Seizure | 6 | 0.683 | 0.135 | 0.674 | 5 |
| L_insula_surfavg | Seizure | 6 | -0.683 | 0.135 | 0.674 | 5 |
| R_insula_surfavg | Seizure | 6 | -0.683 | 0.135 | 0.674 | 5 |
| L_superiorparietal_surfavg | Seizure | 6 | -0.683 | 0.135 | 0.674 | 5 |
| R_posteriorcingulate_surfavg | Seizure | 6 | -0.683 | 0.135 | 0.674 | 5 |
| L_precuneus_surfavg | Seizure | 6 | -0.683 | 0.135 | 0.674 | 5 |
| R_superiorfrontal_surfavg | Seizure | 6 | -0.683 | 0.135 | 0.674 | 5 |
| L_superiorfrontal_surfavg | Seizure | 6 | -0.683 | 0.135 | 0.674 | 5 |
| R_supramarginal_surfavg | Seizure | 6 | -0.683 | 0.135 | 0.674 | 5 |
| R_rostralmiddlefrontal_surfavg | Seizure | 6 | -0.683 | 0.135 | 0.674 | 5 |
| L_lingual_surfavg | Seizure | 6 | -0.683 | 0.135 | 0.674 | 5 |
| L_paracentral_surfavg | Seizure | 6 | -0.683 | 0.135 | 0.674 | 5 |
| R_caudalanteriorcingulate_surfavg | Dysphagia | 6 | -0.655 | 0.158 | 0.792 | 5 |
| R_rostralanteriorcingulate_surfavg | Dysphagia | 6 | -0.655 | 0.158 | 0.792 | 5 |
| Rcaud | Dysphagia | 6 | -0.655 | 0.158 | 0.792 | 5 |
| R_superiorparietal_surfavg | Dysphagia | 6 | 0.655 | 0.158 | 0.792 | 5 |
| L_supramarginal_thickavg | Dysphagia | 6 | -0.655 | 0.158 | 0.792 | 5 |
| L_superiorparietal_thickavg | Dysphagia | 6 | -0.655 | 0.158 | 0.792 | 5 |
| L_lateraloccipital_thickavg | Dysphagia | 6 | 0.655 | 0.158 | 0.792 | 5 |
| L_pericalcarine_surfavg | Dysphagia | 6 | -0.655 | 0.158 | 0.792 | 5 |
| R_precuneus_thickavg | Dysphagia | 6 | -0.655 | 0.158 | 0.792 | 5 |
| L_superiorfrontal_thickavg | Dysphagia | 6 | -0.655 | 0.158 | 0.792 | 5 |
| L_cuneus_surfavg | Dysphagia | 6 | -0.655 | 0.158 | 0.792 | 5 |
| L_rostralmiddlefrontal_thickavg | Dysphagia | 6 | -0.655 | 0.158 | 0.792 | 5 |
| R_caudalanteriorcingulate_surfavg | EEG abnormality | 6 | -0.655 | 0.158 | 0.792 | 5 |
| R_rostralanteriorcingulate_surfavg | EEG abnormality | 6 | -0.655 | 0.158 | 0.792 | 5 |
| Rcaud | EEG abnormality | 6 | -0.655 | 0.158 | 0.792 | 5 |
| R_superiorparietal_surfavg | EEG abnormality | 6 | 0.655 | 0.158 | 0.792 | 5 |
| L_supramarginal_thickavg | EEG abnormality | 6 | -0.655 | 0.158 | 0.792 | 5 |
| L_superiorparietal_thickavg | EEG abnormality | 6 | -0.655 | 0.158 | 0.792 | 5 |
| L_lateraloccipital_thickavg | EEG abnormality | 6 | 0.655 | 0.158 | 0.792 | 5 |
| L_pericalcarine_surfavg | EEG abnormality | 6 | -0.655 | 0.158 | 0.792 | 5 |
| R_precuneus_thickavg | EEG abnormality | 6 | -0.655 | 0.158 | 0.792 | 5 |
| L_superiorfrontal_thickavg | EEG abnormality | 6 | -0.655 | 0.158 | 0.792 | 5 |
| L_cuneus_surfavg | EEG abnormality | 6 | -0.655 | 0.158 | 0.792 | 5 |
| L_rostralmiddlefrontal_thickavg | EEG abnormality | 6 | -0.655 | 0.158 | 0.792 | 5 |
| R_caudalanteriorcingulate_surfavg | Hearing abnormality | 6 | -0.655 | 0.158 | 0.792 | 5 |
| R_rostralanteriorcingulate_surfavg | Hearing abnormality | 6 | -0.655 | 0.158 | 0.792 | 5 |
| Rcaud | Hearing abnormality | 6 | -0.655 | 0.158 | 0.792 | 5 |
| R_superiorparietal_surfavg | Hearing abnormality | 6 | 0.655 | 0.158 | 0.792 | 5 |
| L_supramarginal_thickavg | Hearing abnormality | 6 | -0.655 | 0.158 | 0.792 | 5 |
| L_superiorparietal_thickavg | Hearing abnormality | 6 | -0.655 | 0.158 | 0.792 | 5 |
| L_lateraloccipital_thickavg | Hearing abnormality | 6 | 0.655 | 0.158 | 0.792 | 5 |
| L_pericalcarine_surfavg | Hearing abnormality | 6 | -0.655 | 0.158 | 0.792 | 5 |
| R_precuneus_thickavg | Hearing abnormality | 6 | -0.655 | 0.158 | 0.792 | 5 |
| L_superiorfrontal_thickavg | Hearing abnormality | 6 | -0.655 | 0.158 | 0.792 | 5 |
| L_cuneus_surfavg | Hearing abnormality | 6 | -0.655 | 0.158 | 0.792 | 5 |
| L_rostralmiddlefrontal_thickavg | Hearing abnormality | 6 | -0.655 | 0.158 | 0.792 | 5 |
| R_rostralmiddlefrontal_thickavg | Hyperkinetic movements | 6 | -0.63 | 0.18 | 0.9 | 5 |
| L_parsopercularis_thickavg | Dysphagia | 5 | 0.707 | 0.182 | 0.908 | 5 |
| L_parsorbitalis_thickavg | Dysphagia | 5 | 0.707 | 0.182 | 0.908 | 5 |
| L_parsopercularis_thickavg | EEG abnormality | 5 | 0.707 | 0.182 | 0.908 | 5 |
| L_parsorbitalis_thickavg | EEG abnormality | 5 | 0.707 | 0.182 | 0.908 | 5 |
| L_parsopercularis_thickavg | Hearing abnormality | 5 | 0.707 | 0.182 | 0.908 | 5 |
| L_parsorbitalis_thickavg | Hearing abnormality | 5 | 0.707 | 0.182 | 0.908 | 5 |
| L_superiorparietal_thickavg | Gait disturbance | 6 | -0.621 | 0.188 | 0.941 | 5 |
| R_lateralorbitofrontal_surfavg | Gait disturbance | 6 | -0.621 | 0.188 | 0.941 | 5 |
| L_lingual_surfavg | Gait disturbance | 6 | -0.621 | 0.188 | 0.941 | 5 |
| L_pericalcarine_surfavg | Gait disturbance | 6 | -0.621 | 0.188 | 0.941 | 5 |
| R_rostralanteriorcingulate_surfavg | Generalized hypotonia | 6 | -0.621 | 0.188 | 0.941 | 5 |
| L_lingual_surfavg | Hyperkinetic movements | 6 | -0.621 | 0.188 | 0.941 | 5 |
| L_pericalcarine_surfavg | Hyperkinetic movements | 6 | -0.621 | 0.188 | 0.941 | 5 |
| L_superiorparietal_thickavg | Sleep abnormality | 6 | -0.621 | 0.188 | 0.941 | 5 |
| R_lateralorbitofrontal_surfavg | Sleep abnormality | 6 | -0.621 | 0.188 | 0.941 | 5 |
| L_lingual_surfavg | Sleep abnormality | 6 | -0.621 | 0.188 | 0.941 | 5 |
| L_pericalcarine_surfavg | Sleep abnormality | 6 | -0.621 | 0.188 | 0.941 | 5 |
| L_caudalanteriorcingulate_surfavg | Gait disturbance | 6 | 0.621 | 0.188 | 0.941 | 5 |
| L_caudalmiddlefrontal_surfavg | Gait disturbance | 6 | 0.621 | 0.188 | 0.941 | 5 |
| L_lateraloccipital_thickavg | Gait disturbance | 6 | 0.621 | 0.188 | 0.941 | 5 |
| R_caudalanteriorcingulate_thickavg | Gait disturbance | 6 | 0.621 | 0.188 | 0.941 | 5 |
| L_posteriorcingulate_surfavg | Generalized hypotonia | 6 | 0.621 | 0.188 | 0.941 | 5 |
| L_postcentral_surfavg | Generalized hypotonia | 6 | 0.621 | 0.188 | 0.941 | 5 |
| R_superiorparietal_surfavg | Generalized hypotonia | 6 | 0.621 | 0.188 | 0.941 | 5 |
| L_supramarginal_surfavg | Generalized hypotonia | 6 | 0.621 | 0.188 | 0.941 | 5 |
| L_cuneus_thickavg | Hyperkinetic movements | 6 | 0.621 | 0.188 | 0.941 | 5 |
| L_lateraloccipital_thickavg | Hyperkinetic movements | 6 | 0.621 | 0.188 | 0.941 | 5 |
| L_caudalanteriorcingulate_surfavg | Sleep abnormality | 6 | 0.621 | 0.188 | 0.941 | 5 |
| L_caudalmiddlefrontal_surfavg | Sleep abnormality | 6 | 0.621 | 0.188 | 0.941 | 5 |
| L_lateraloccipital_thickavg | Sleep abnormality | 6 | 0.621 | 0.188 | 0.941 | 5 |
| R_caudalanteriorcingulate_thickavg | Sleep abnormality | 6 | 0.621 | 0.188 | 0.941 | 5 |

**Supplementary Table 4.** **Complete clinical associations in the older patient group.** Among the regions where the mean Z-score differed from 0 with *p*<.10, we examined associations (presence or absence) with 9 clinical measures. For each pair, the number of patients included, Spearman ρ (rho), raw *p*-values, adjusted *p*-values using the Li & Ji method, and the effective number of tests calculated by the Li & Ji method are reported.

| **Region** | **Clinical_measure** | **n_pairs** | **ρ** | **p_value** | **p_adjusted_li_ji** | **m_eff** |
| --- | --- | --- | --- | --- | --- | --- |
| Rthal | Dysphagia | 7 | -0.866 | 0.012 | 0.094 | 8 |
| Lthal | Dysphagia | 7 | -0.866 | 0.012 | 0.094 | 8 |
| Rthal | EEG_abnormality | 7 | -0.866 | 0.012 | 0.094 | 8 |
| Lthal | EEG_abnormality | 7 | -0.866 | 0.012 | 0.094 | 8 |
| Rthal | Gait_disturbance | 7 | -0.866 | 0.012 | 0.094 | 8 |
| Lthal | Gait_disturbance | 7 | -0.866 | 0.012 | 0.094 | 8 |
| L_temporalpole_surfavg | Hearing_abnormality | 7 | -0.866 | 0.012 | 0.094 | 8 |
| L_lateraloccipital_thickavg | Hearing_abnormality | 7 | -0.866 | 0.012 | 0.094 | 8 |
| L_lateraloccipital_surfavg | Sleep_abnormality | 7 | -0.866 | 0.012 | 0.094 | 8 |
| L_isthmuscingulate_thickavg | Hyperkinetic_movements | 7 | 0.866 | 0.012 | 0.094 | 8 |
| R_insula_surfavg | Seizure | 7 | 0.866 | 0.012 | 0.094 | 8 |
| Lpal | Sleep_abnormality | 7 | 0.866 | 0.012 | 0.094 | 8 |
| Rhippo | Sleep_abnormality | 7 | 0.866 | 0.012 | 0.094 | 8 |
| R_supramargil_surfavg | Sleep_abnormality | 7 | 0.866 | 0.012 | 0.094 | 8 |
| Lthal | Tremor | 7 | -0.791 | 0.034 | 0.275 | 8 |
| L_superiorfrontal_surfavg | Tremor | 7 | 0.791 | 0.034 | 0.275 | 8 |
| R_insula_surfavg | Tremor | 7 | 0.791 | 0.034 | 0.275 | 8 |
| R_fusiform_thickavg | Tremor | 7 | -0.791 | 0.034 | 0.275 | 8 |
| L_rostralmiddlefrontal_surfavg | Tremor | 7 | 0.791 | 0.034 | 0.275 | 8 |
| L_lateraloccipital_surfavg | Dysphagia | 7 | -0.722 | 0.067 | 0.537 | 8 |
| L_lateraloccipital_surfavg | EEG_abnormality | 7 | -0.722 | 0.067 | 0.537 | 8 |
| L_lateraloccipital_surfavg | Gait_disturbance | 7 | -0.722 | 0.067 | 0.537 | 8 |
| L_rostralanteriorcingulate_thickavg | Hearing_abnormality | 7 | -0.722 | 0.067 | 0.537 | 8 |
| R_rostralanteriorcingulate_surfavg | Hyperkinetic_movements | 7 | -0.722 | 0.067 | 0.537 | 8 |
| L_cuneus_thickavg | Seizure | 7 | -0.722 | 0.067 | 0.537 | 8 |
| R_medialorbitofrontal_surfavg | Seizure | 7 | -0.722 | 0.067 | 0.537 | 8 |
| Rthal | Sleep_abnormality | 7 | -0.722 | 0.067 | 0.537 | 8 |
| L_transversetemporal_thickavg | Hearing_abnormality | 7 | 0.722 | 0.067 | 0.537 | 8 |
| L_isthmuscingulate_thickavg | Hearing_abnormality | 7 | 0.722 | 0.067 | 0.537 | 8 |
| L_superiorfrontal_surfavg | Seizure | 7 | 0.722 | 0.067 | 0.537 | 8 |
| L_rostralmiddlefrontal_surfavg | Seizure | 7 | 0.722 | 0.067 | 0.537 | 8 |
| Lhippo | Sleep_abnormality | 7 | 0.722 | 0.067 | 0.537 | 8 |
| Lput | Generalized_hypotonia | 7 | -0.612 | 0.144 | 1 | 8 |
| Laccumb | Generalized_hypotonia | 7 | 0.612 | 0.144 | 1 | 8 |
| L_frontalpole_surfavg | Generalized_hypotonia | 7 | 0.612 | 0.144 | 1 | 8 |
| L_rostralanteriorcingulate_thickavg | Generalized_hypotonia | 7 | 0.612 | 0.144 | 1 | 8 |
| R_postcentral_surfavg | Generalized_hypotonia | 7 | -0.612 | 0.144 | 1 | 8 |
| L_postcentral_surfavg | Generalized_hypotonia | 7 | 0.612 | 0.144 | 1 | 8 |
| L_fusiform_thickavg | Generalized_hypotonia | 7 | 0.612 | 0.144 | 1 | 8 |
| R_medialorbitofrontal_surfavg | Generalized_hypotonia | 7 | 0.612 | 0.144 | 1 | 8 |
| L_insula_thickavg | Generalized_hypotonia | 7 | -0.612 | 0.144 | 1 | 8 |
| R_fusiform_surfavg | Tremor | 7 | 0.632 | 0.127 | 1 | 8 |
| L_lingual_thickavg | Tremor | 7 | -0.632 | 0.127 | 1 | 8 |
| L_insula_thickavg | Tremor | 7 | -0.632 | 0.127 | 1 | 8 |
| L_transversetemporal_surfavg | Tremor | 7 | -0.632 | 0.127 | 1 | 8 |
| L_postcentral_thickavg | Tremor | 7 | 0.632 | 0.127 | 1 | 8 |
